# A Blood Transcriptomic Resource for ALS Highlights Disease-Associated Signatures and Alternative Splicing Events

**DOI:** 10.1101/2025.07.28.25332310

**Authors:** Cheong W. Wong, Laura Ziser, Lachlan Sparke, Ruolan Zhao, Anna Freydenzon, Solal Chauquet, Robert Henderson, Shyuan Ngo, Leanne Wallace, Naomi R Wray, Anjali K Henders, Pamela McCombe, Allan McRae, Fleur C Garton

## Abstract

Amyotrophic lateral sclerosis (ALS) is a neurogenerative disease resulting from progressive degeneration of motor neurons leading to systemic consequences. Despite being the most common motor neuron disease, with increasing global prevalence, limited treatment options exist. Emerging evidence from genetic studies and pathology analyses implicates RNA dysregulation in ALS pathogenesis, however, deep, comprehensive RNA sequencing studies have not been carried out.

Here, we analysed >240 ALS and control whole blood transcriptome samples. Cross-sectional (N_cases_=121, N_controls_=53) and longitudinal (N_observations_=103) cohorts supported complementary expression analyses of disease mechanisms across disease stages. Both short (N=241) and long-read (N=16) technologies were utilised to discover splicing changes. Total RNA was extracted from PAXgene whole blood RNA tubes before libraries (Illumina Stranded Total RNA RiboZero Plus) were prepared and sequenced (∼50M PE reads per sample). Long-read sequencing was performed using the Mas-Seq protocol with Kinnex full-length RNA prep kit and sequenced (PacBio Revio platform, 10M reads per sample) for full-length transcripts.

Case-control cohort analyses identified 50 significantly differentially expressed genes, with pathway analyses implicating RNA processing and immune system regulation. Findings were corroborated using existing ALS RNAseq datasets from blood (correlation >0.4), iPSC-MN and post-mortem tissues. Alternative splicing (AS) analyses (LeafCutter) identified 62 clusters. Within-case analyses involved ALS cases with multiple (2-4) visits, detected 144 genes associated with disability progression over time. The long-read sequencing (N_cases_=8, N_controls_=8) provided novel discovery insights, in particular in the HLA region.

This comprehensive blood-based transcriptomic dataset reveals both known and novel disease mechanisms in ALS, offering valuable insights that could inform future research and therapeutic development. The results of this study may inform and refine the prioritization of candidate genes and loci in future ALS research.

## Introduction

Amyotrophic lateral sclerosis (ALS) is a neurogenerative disease caused by progressive loss of the motor neurons in the central nervous system (CNS) [1]. System-wide impacts are felt, with muscle weakness, loss of motor skills, communication and daily task difficulty with compromised respiratory function contributing to early death [2]. There is no current therapeutic or intervention can effectively cure or reverse the damage of the disease. Nonetheless, understanding on the pathogenesis of ALS has been expanding with both Mendelian genes and large genome-wide association studies contributing to this knowledge on disease risk and progression [3–6]. There is growing evidence that RNA dysregulation may play a central role in ALS pathogenesis. Several genetic risk loci (i.e. *ATXN2*) [7–9], causal genes (i.e. *C9orf72* [10], *TARDBP* [11, 12], *FUS* [13]) and pathological hallmarks (TDP-43 inclusions, [14, 15]) are all involved in RNA processing, highlighting its importance for motor neuron health [16].

RNA sequencing (RNAseq) captures not only gene expression changes but also isoform splicing alterations– both of which are relevant to ALS pathogenesis [7, 8]. Blood is a minimally invasive, easily accessible tissue making it practical for large-scale, longitudinal sampling. This supports the investigation of systemic mechanisms underlying the body’s response to disease [17–19]. With limited studies being carried out in this space the sporadic ALS disease transcriptome remains largely unexplored.

To tackle this resource gap, here we contribute a large ALS transcriptome resource to the community with relatively deep sequenced, short and long read data (with matched clinical and genetic SNP-array data) to discover mechanisms of ALS and disease risk. In this blood-based investigation, we tested two main hypotheses: 1) ALS cases would show altered isoforms and transcript expression compared to controls, and 2) a proportion of novel events would be detected in the transcriptome of ALS cases. We find evidence to support blood as providing relevant ALS disease signals, with both similarities and differences (as expected) with post-mortem/iPSC-MN models, as well as discovering new unique signals that could indeed be relevant to the disease process.

## Methods

### Sample collection and processing

All of the subjects were recruited from Brisbane, Australia between 2012–2022 with approval from the local Human Research Ethics Committees (HREC Metro North Health EC00172 (2006/047) and the University of Queensland (2021/HE002682)). Cases were clinically diagnosed ALS patients according to the El Escorial criteria and controls were similarly aged (partners, carers or community controls) free from known neurodegenerative diseases. The initial dataset consisted of 252 whole blood samples collected in PAXgene RNA blood tubes (stored at −80°C until use) from 178 individuals (N_cases_=188, N_controls_= 64). Briefly, 2.5mls of whole blood was thawed and RNA was extracted using the QIAcube robot with the PAXgene Blood miRNA Kit (mean RIN value = 7.98 ± 1.10 SD). Libraries were prepared using the Illumina Stranded Total RNA RiboZero Plus kit at the University of Queensland (Brisbane). After quality control checks, sequencing was generated on the DNBSEQ-T7 platform (150bp, ∼50M paired-end) at the South Australian Health and Medical Research Institute Sequencing facility (**Figure 1**).

**Figure 1.**
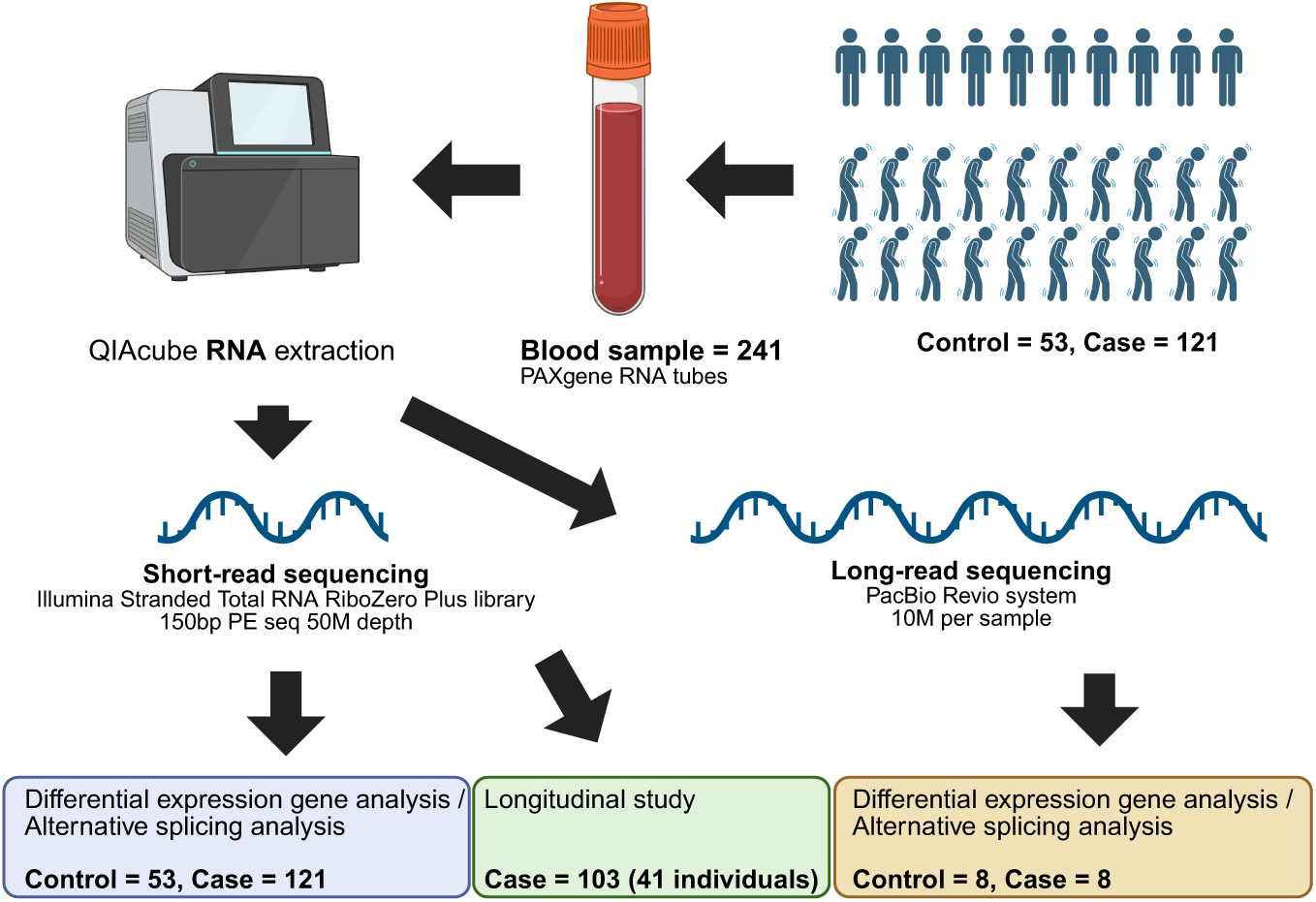
Schematic of the experiment design. Starting from the top right; a total of 241 blood samples were collected from 174 individuals in PAXgene tubes before being extracted using the QIAcube. Extracted RNA was then prepared for short- and long-read RNA sequencing. To carry out short-read sequencing, Illumina stranded total RNA (RiboZero Plus) libraries were prepared (∼50M paired-end reads/sample) this comprised of a cross-sectional cohort (ALS cases = 121, controls = 53) and a longitudinal cohort (103 observations from repeat visits). A subset of samples (n = 16; 8 cases, 8 controls) underwent full-length transcript profiling using the MAS-Seq protocol with the Kinnex full-length RNA prep kit (PacBio Revio platform, ∼10M reads/sample). Analyses included differential expression and alternative splicing analyses with findings validated using external RNA-seq datasets.

### RNA sequencing quality control and data-analysis

Read quality was assessed using FastQC (v0.11.9) and inspected using MultiQC (v1.14-foss-2022a). Low quality reads (mean Phred score < 15) were removed and adapter sequences were trimmed using *fastp* [20]. Sequences were aligned with STAR [21] and mapped to the GRCh38 genome reference sequence. Annotated transcripts (gencode.v43) were used to generate BAM files for gene count summarization. Read counts were summarized using featureCounts [22] putting “gene” as the feature type (v4.4.3). The gene expression matrix of the full dataset 241 samples was subjected to further quality control in R (v4.4.3). All individual samples had a raw total gene expression count of >20,000,000 except one (slightly below the threshold, N = 18,594,156). Two-hundred and forty-one samples were retained for further quality checking such as subgroup identification. The processed dataset was investigated by 1) differential gene expression (DE) analysis in cases and controls (contained only one visit (random selection) if multiple existed) (**Table 1**) and within-case analyses of disease progression (case only, **Supplementary Table 1**).

**Table 1.** Demographic and clinical data of the differential gene expression sample.

|  | **Control** | **Case** | **Total** | **P-value<sup>#</sup>/ Note** |
| --- | --- | --- | --- | --- |
| **Number** | 53 (30.5%) | 121 (69.5%) | 174 | >0.01<br>Total Male vs<br>Female |
| **Male** | 29 (26.6%) | 80 (73.4%) | 109<br>(62.6%) |  |
| **Female** | 24 (36.9%) | 41 (63.1%) | 65<br>(37.4%) |  |
| **Age at collection**<br>**(years)** | 59.9 ±10.3 | 61.9 ±9.6 | NA | 0.24 |
| **Age at onset (years)** | NA | 59.6 ±9.8 | NA | Range ~ 32 – 96 |
| **Family Hx of ALS**<br>**and/or FTD** | 0 | 16 | 16 | - |
| ***C9orf72* HRE**<br>**positive** | 0 | 2 | NA | - |
| ***SOD1* or *TARDBP***<br>**positive** | 0 | 1 | 1 | - |
| **Time between**<br>**collection and onset**<br>**(months)** | NA | 28.7 ± 22.5 | NA | - |

Table legend: ± denotes standard deviation, P-value = t-test, HRE= Hexonucleotide repeat expansion carrier =>30 repeats. *SOD1/TARDBP* screening based on likely pathogenic/pathogenic variants in custom SNP array (contains most frequent variants), Hx= History of family member having dementia / ALS.

### Differential gene expression analysis

The differential gene expression (DE) dataset consisted of initially 178 blood samples. Two samples were excluded due to sex/gender mismatch, identified by plotting *XIST* (female) and *ZFY* (male specific) gene expression (log_2_(CPM+1)); **Supplementary Figure 1**) and an additional two were removed due to missing age, resulting in a final dataset of 174 samples (N_cases_= 121, N_controls_ = 53, **Table 1**).

Principal components analysis (PCA) showed clear separation between males and female samples (**Supplementary Figure 2**) highlighting sex as an important covariate within our model. No clustering based on batch or other known technical or biological factors were observed (**Supplementary Figure 3**).

Genes with a mean count ≤10 across all samples were filtered out, resulting in 19,521 being retained for analysis. The DE analysis was performed using *DESeq2* [23] comparing cases to controls while adjusting for age and sex (design = ∼ Age + Sex + ALS_status). Genes with a Benjamini-Hochberg adjusted p-value ≤0.05 for ALS status were identified as DE genes in this analysis.

### Pathway, Gene set and tissue enrichment analyses

The DE genes using Ensembl IDs were extracted for pathway enrichment analysis performed using Metascape (v3.5.20240901). The list of identified DE genes was matched against the default set of databases (GO biological process [24], KEGG pathways [25], Reactome gene sets [26], CORUM complexes [27], and canonical pathways from MSigDB [28]). Pathways that were significantly enriched in at least three databases, a p-value > 0.01, and minimum enrichment of 1.5 factor were indicated as significant.

To explore biological enriched pathways in the DE dataset, gene set enrichment analysis (GSEA) was performed to analysis the full set of DE results. R packages *fgsea* [29] was used for this GSEA analysis. All genes in the DE result were ranked based on the enrichment. The ranked gene list was compared to gene sets in the Molecular Signatures Database (MSigDB) [30]. Minimum size of gene sets in the analysis was kept at 15 and visualized in R.

Tissue specificity enrichment analyses were carried out using the significant genes vs. all genes detected in the GTEx database (54 human tissues, v8). Normalised expression values were used (zero-mean, log_2_ expression TPM). Differentially Expressed Gene Sets were defined as genes with a p-value of ≤ 0.05 after Bonferroni correction (54 tests) and absolute log fold change ≥ 0.58. Enrichment was tested using the hypergeometric test, with background genes defined as those included in the analysis and with an average expression value > 1 TPM.

### Longitudinal cohort and within-case analyses

The longitudinal cohort dataset consisted of 41 cases (male = 32, female = 9) and 103 blood collections, all processed together using consistent methods as described above. Each collection had a matched clinical score on the ALS Functional Rating Scale–Revised (ALSFRS-R), which ranges from 48 (no disability) to 0 (severe disability). Of the 41 individuals, 23 had two collections, 15 had three, and 3 had four, with an average interval of 8.6 months. Date of disease onset was based on first reported symptoms at their initial visit.

Rate of progression for each individual was calculated as:

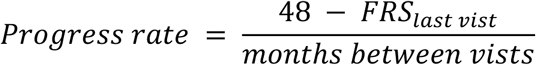

The average progress rate was 0.55 (*SD* 0.46); most individuals progressed at a rate of ≤ 1 point/month, while two cases progressed faster >2 points/month (**Supplementary Figures 4 and 51**).

We tested three models using the R package *variancePartition* [31], applying linear mixed models to transformed counts (log_2_-counts per million (logCPM) (*voomWithDreamWeights* function)). As above, genes that had a mean count >10 across samples were excluded, leaving ∼19,260 genes per analysis. Repeated measures were accounted for with a random intercept per individual.

Our primary analysis tested whether ALS disease severity (ALSFRS-R) was associated with gene expression across timepoints.

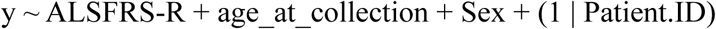

Secondly, we tested a change (delta-delta) model, asking whether within-individual changes in gene expression were associated with changes in ALSFRS over time.

The residuals:

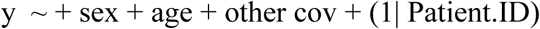

were used to model:

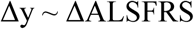

using “pseudo independent pairs” (e.g. visit 1-2, 1-3, 2-3, for those with ≥3 visits).

Finally, we ran a cross-sectional case-only model, testing whether gene expression correlated with ALSFRS-R at each timepoint (including both single- and multi-visit samples):

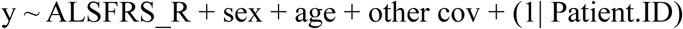

For all analyses, p-values were adjusted with the Benjamini-Hochberg method. If variance partitioning was applied, the same R package with function *fitExtractVarPartModel*, was used on the normalised count matrix, modelling all categorical variables (e.g., sex) as random effects as required by the package.

### Alternative splicing analysis

The dataset used in this alternative splicing analysis was the same as the DE dataset (174 samples (N_cases_= 121, N_controls_ = 53)). Alternative splicing events were quantified in controls and cases using LeafCutter (v0.2.7) [32]. Intron excision was detected based on split mapped reads of the RNAseq data. Introns that shared common splicing junctions were grouped into clusters. Alternative splicing events were estimated by the quantities of reads mapped to the introns in the clusters. Clusters were identified based on minimum 50 split reads and maximum intron size of 500kb (default parameters). Introns differentially spliced among controls and cases were identified with sex and age as covariates. Results were sorted and visualized on R package Leafviz. To prioritize potential genes of interest for future analysis, results were intersected with genes with a high probability of loss-of-function intolerant score pLI score ≥ 0.9 (generated from the Genome Aggregation Database (gnomAD)) [33], a list of TDP-43 binding partners impacted by knock-down [34], and ALS/or similar disease genes (causal, mapped risk GWAS threshold = 5e-7) [4, 35].

### Long-read sequencing sample processing and analyses

A set of 16 samples (N_cases_=8, N_controls_=8, mean age 60 ± 7.69 SD) in the DE dataset was selected for long-read sequencing. Selection was based on high-quality RNA remaining from the earlier extractions for second library preparation (average RIN value 8.9 ± 0.31), enrichment of gene carriers (1x *TARDBP* and 2x *C9orf72*) and sex/age matching cases/controls. Remaining case individuals within the cohort displayed ALS symptoms without known ALS variants. The libraries were prepared using the MAS-Seq protocol with Kinnex full-length RNA kit (PacBio) and primers in the Iso-Seq® Express 2.0 Kit. Multiplexed samples were sequenced with PacBio Revio system at 10M reads per sample at the University of Queensland, Brisbane. Generated Hi-Fi reads were converted to BAM files and further processed by FLAIR [36]. Reads were aligned and transcripts annotated against the GRCh38 reference genome. Differential expression and alternative splicing analyses were conducted between cases and controls. Filtered gene count matrix from FLAIR was extracted and analysed using the same *DEseq2* configurations as the short-read data.

### Cell type deconvolution

We estimated the proportion of cell types of the bulk whole-blood, short-read RNA-seq DE dataset using CIBERSORTx [37] with LM22 (containing 22 subtypes of immune cells) used as the signature matrix. Transcript abundance in transcripts per million (TPM) was estimated by Salmon [38] and collated by tximport [39]. Quantile normalization was disabled, and 100 permutations were used to assess for statistical significance. Estimated subtype proportions were summed up (B cells = B cells naive + B cells memory, T cells = T cells CD8 + T cells CD4 naive + T cells CD4 memory resting + T cells CD4 memory activated + T cells follicular helper + T cells regulatory + T cells gamma delta, NK cells = NK cells resting + NK cells activated). Kolmogorov–Smirnov test and Wilcoxon signed-rank test were used the statistical differences in estimated cell type proportions between controls and cases.

### Gene/transcript validation in external blood, iPSC-MN and postmortem RNAseq data

To validate the results in this study (UQ), external publicly available blood, iPSC-MN (induced pluripotent stem cell derived motor neurons) and postmortem RNAseq data were analysed. The largest available blood RNAseq dataset was downloaded from NCBI GEO (accession GSE234297, [40]) and the QC and DE pipeline described above was applied with one exception; the filter removing samples with total read count ≤ 20,000,000 was not used due to the dataset’s lower sequencing coverage (only two samples would have remained). Wald statistic values generated from the DE analyses in both the UQ and GSE234297 results were extracted and compared. To assess dataset similarity, raw count matrices from both datasets were combined (processed with the same pipeline) and pairwise Pearson correlations were computed to generate a correlation matrix visualised with a heatmap. Identified DE genes in both the UQ result and GSE234297 were intersected with the original DE result (called original here to avoid confusion) reported from [40].

The largest meta-analysed iPSC-MN and postmortem tissue RNAseq analysis was taken from Ziff et al. [41]. Differentially expressed gene results including pan-ALS (N_sample_ = 429) and post-mortem spinal cord tissue (N_sample_ = 271) were downloaded [41] and intersected in R with the blood DE result from the UQ dataset. Only identified DE genes (FDR < 0.05) were included in this meta-analysis

## Results

### Detection of Differentially Expressed gene in blood samples from ALS patients

Whole blood transcriptome differential expression analysis in ALS cases vs controls (N_cases_=121 and N_controls_=53) identified 50 DE genes (FDR ≤ 0.05) (**Figure 2a**, **Table 2, Supplementary Table 2**). The log_2_ fold changes were modest (range 0.06-2.02) with a mix of both upregulated (n=24) and downregulated gene expression changes (n=26).

**Figure 2.**
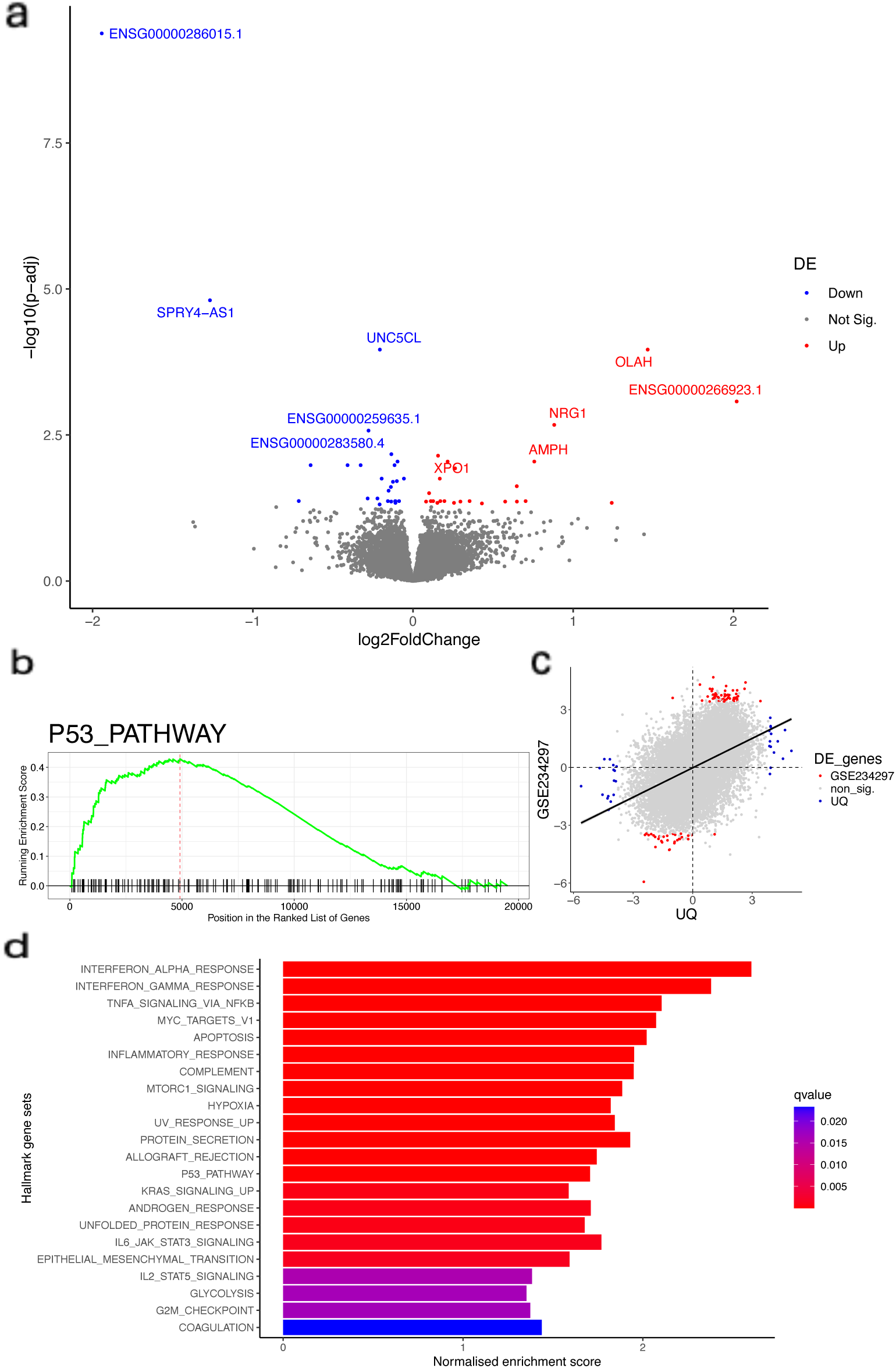
**a)** Differential expression in ALS identified 50 DE genes (FDR ≤ 0.05) (N_cases_=121, N_controls_=53) with the top hit (downregulated >1.94 log_2_FC) typically found in brain (ENSG00000286015). Red dots indicate the significantly upregulated genes and blue, significantly down regulated. The top 10 genes are labelled by their symbols/Ensembl IDs **b)** Enrichment analyses of DE genes detected 22 enriched pathways **(d)**, with the p53 pathway shown in detail. This visualises the relative positions of p53 pathway genes in the DE analysis. Genes were ranked according to their adjusted p-values and their direction of fold changes (left – up regulated, right – down regulated). From left to right every encounter of a gene involved in the p53 pathway moves the green line up, vice versa. Dotted red line (the peak of the green line) represents the enrichment score. The peak is relative to the left of the centre, indicating the identified DE genes that were involved in this pathway were mostly upregulated. **c)** Positive correlation of DE results with a second ALS RNAseq study (GSE234297) (N_cases_=96, N_controls_=48) despite lower coverage/differing library kit preparation (**Supplementary Note**). Wald statistic values of corresponding genes were plotted. Red dot represents a gene that was identified as a DE gene in both studies, green only in this study, blue only in GSE234297. *β*-coefficient of the regression line = 0.42 (p-value < 2.2e-16). R^2^ = 0.21. Sixty-seven percent of the genes were in I and III quadrant.

**Table 2.**
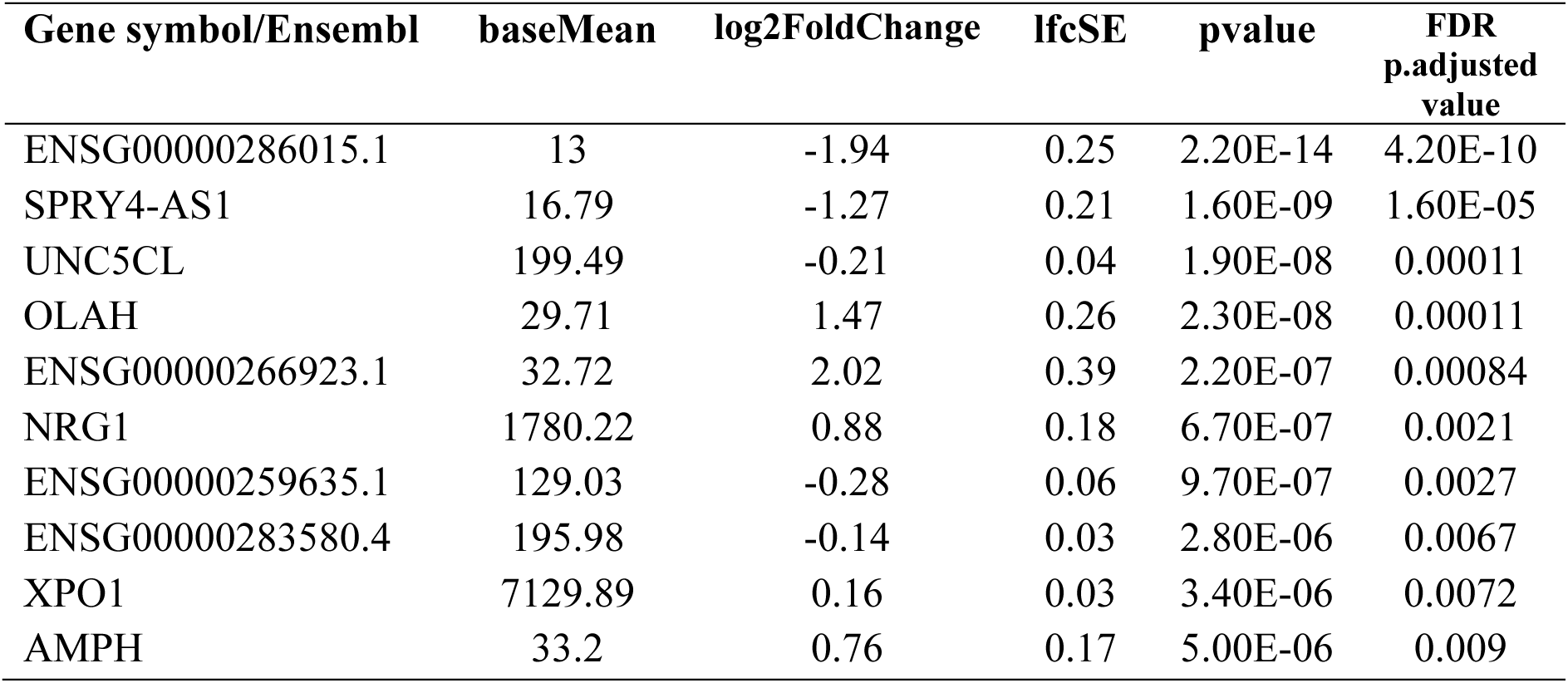
Top 10 representative results of 50 significant differentially expressed genes (N_cases_=121, N_controls_=53)

Although blood is considered a peripheral tissue to ALS disease, we next explored whether the DE expressed detected in blood might reflect any particular tissue enrichment expression pattern. Expression annotations from human tissues in GTEx [42] and the Human Protein Atlas [43] were summarised (**Supplementary Figure 6**). DE genes did not show any specific enrichment overall, with six uncharacterised [42]. Notably, the top DE gene (ENSG00000286015, adjusted p-value=4.20e-10), which was downregulated in ALS cases (log_2_FC= −1.94), has previously only been detected in brain tissue [43]. More broadly, no enrichment pattern was observed across different tissues (**Supplementary Figure 6**).

### Biological and functional analysis using enrichment pathway analysis and GSEA

To gain insight into the biological functions of the DEGs, we performed enrichment pathway analysis and gene set enrichment analysis (GSEA). Pathways that were enriched in the gene list could potentially reflect what were the biological responses to ALS in the body. The GSEA identified 22 significant pathways (**Supplementary Table 3**), some of the pathways (e.g. Hallmark Interferon Alpha Response, Hallmark Interferon Gamma Response, etc.) were related to the immune system. Pathways that were related to cell proliferation and death (e.g. Hallmark Apoptosis, Hallmark MYC Targets V1 and Hallmark TNFA signalling via NFKB) and the p53 pathway (NES = 1.71, FDR 5.94e-05) were also identified (**Figure 2b**). Metascape enrichment pathway analysis was used as a complementary approach, detecting five enriched pathways, again encompassing pathways related to immune responses (e.g. GO:0031349) and RNA processing (e.g. R-HSA-8953854) (**Supplementary Figure 7**).

### Longitudinal cohort and within-case analyses

To assess changes in gene expression over the course of disease progression, we analysed 103 observations from 41 cases. Although the rate of decline varied between individuals, nearly all timepoints showed a decrease in ALS Functional Rating Score-Revised (ALSFRS-R) over time (**Figure 3a**, **Supplementary Figure 4 and 5**).

**Figure 3.**
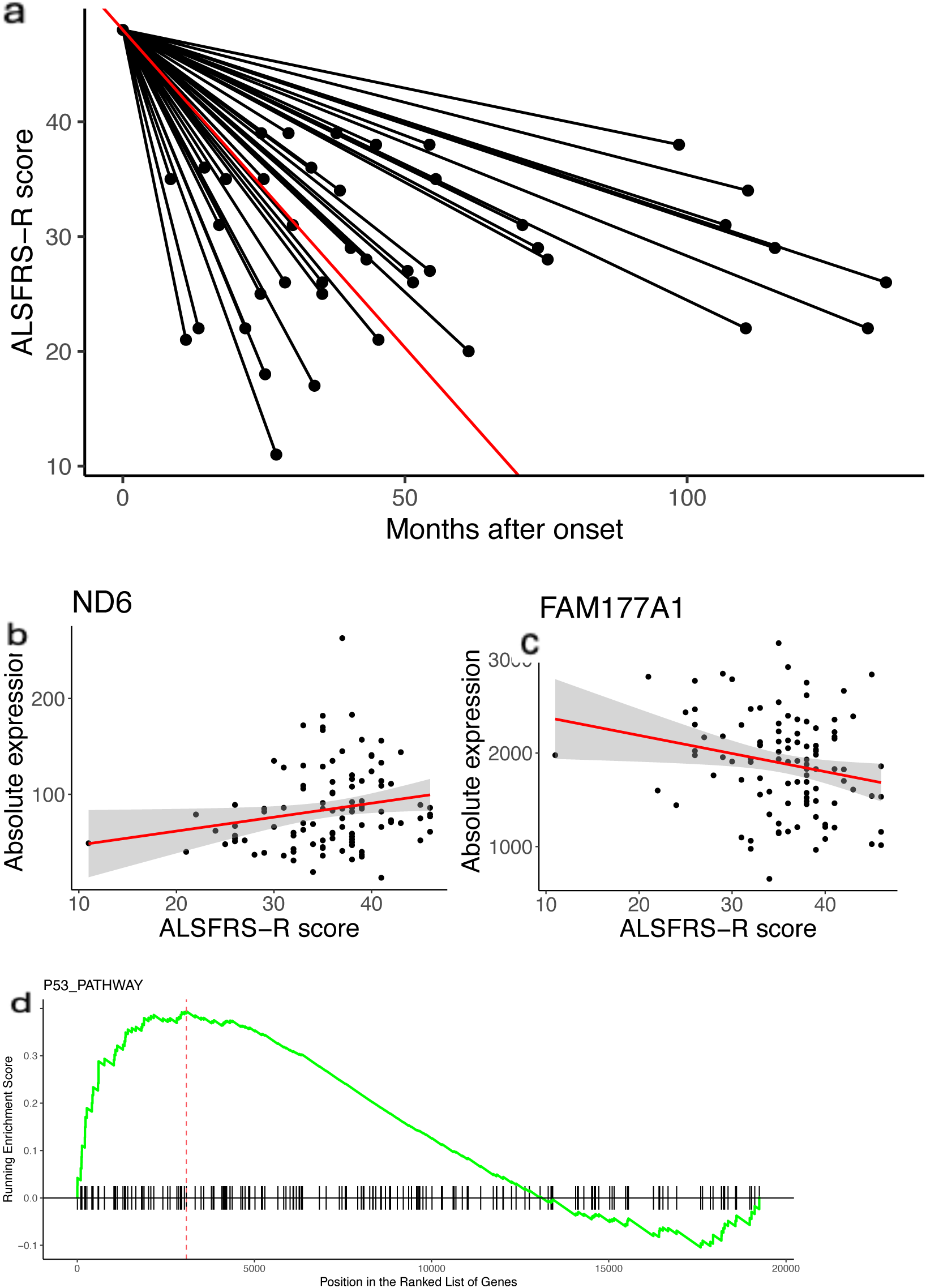
**a)** Progression rates of ALS in the longitudinal cohort (N_cases_=41, N_observations_=103) calculated by measuring the change in ALSFRS between the last visit and the onset and divided by months). The red line is the average progression rate (−0.55,*SD* ±0.46), the black is each individual case. **b)** Linear model of *ND6* expression depending on ALSFRS-R with age and sex as covariates (*β* = 0.06, adjusted p-value = 0.002). **c)** Linear model of *FAM177A1* expression depending on ALSFRS-R with age and sex as covariates (β = −0.011, adjusted p-value = 0.004). **d)** Enrichment analyses of longitudinal study detected 36 enriched pathways, with the p53 pathway shown (previously detected in ALS iPSC-MN/PM study). This visualises the relative positions of p53 pathway genes in the DE analysis. Genes were ranked according to their adjusted p-values and their direction of fold changes (left – up regulated, right – down regulated). From left to right every encounter of a gene involved in the p53 pathway moves the green line up, vice versa. Dotted red line (the peak of the green line) represents the enrichment score. The peak is relative to the left of the centre, indicating the identified DE genes that were involved in this pathway were mostly upregulated.

The primary, within-case gene-expression analysis identified 144 genes significantly associated with disease progression over time, with just one gene that overlapped with the case/control DE analysis (*LYPD2*) (**Table 3**, **Supplementary Table 4**). The top up-regulated gene in ALS cases (*ND6*: *β* = 0.06, adjusted p-value = 0.002, **Figure 3b**) has a more enriched expression in heart and skeletal muscle [43]. The top down-regulated gene associated with progression in ALS cases (*FAM177A1*: *β* = −0.01, adjusted p-value = 0.004, **Figure 3c**) had low tissue specificity expression pattern [43]. The average absolute *β*-coefficient was 0.032 ± 0.015. A quantile-quantile (Q-Q) plot revealed p-value inflation (λ= 1.91) (**Supplementary Figure 8**), which is not unexpected for longitudinal analyses [44]. Moreover, most of the variance was observed to come from between individuals or be unexplained (**Supplementary Figure 9**).

**Table 3.**
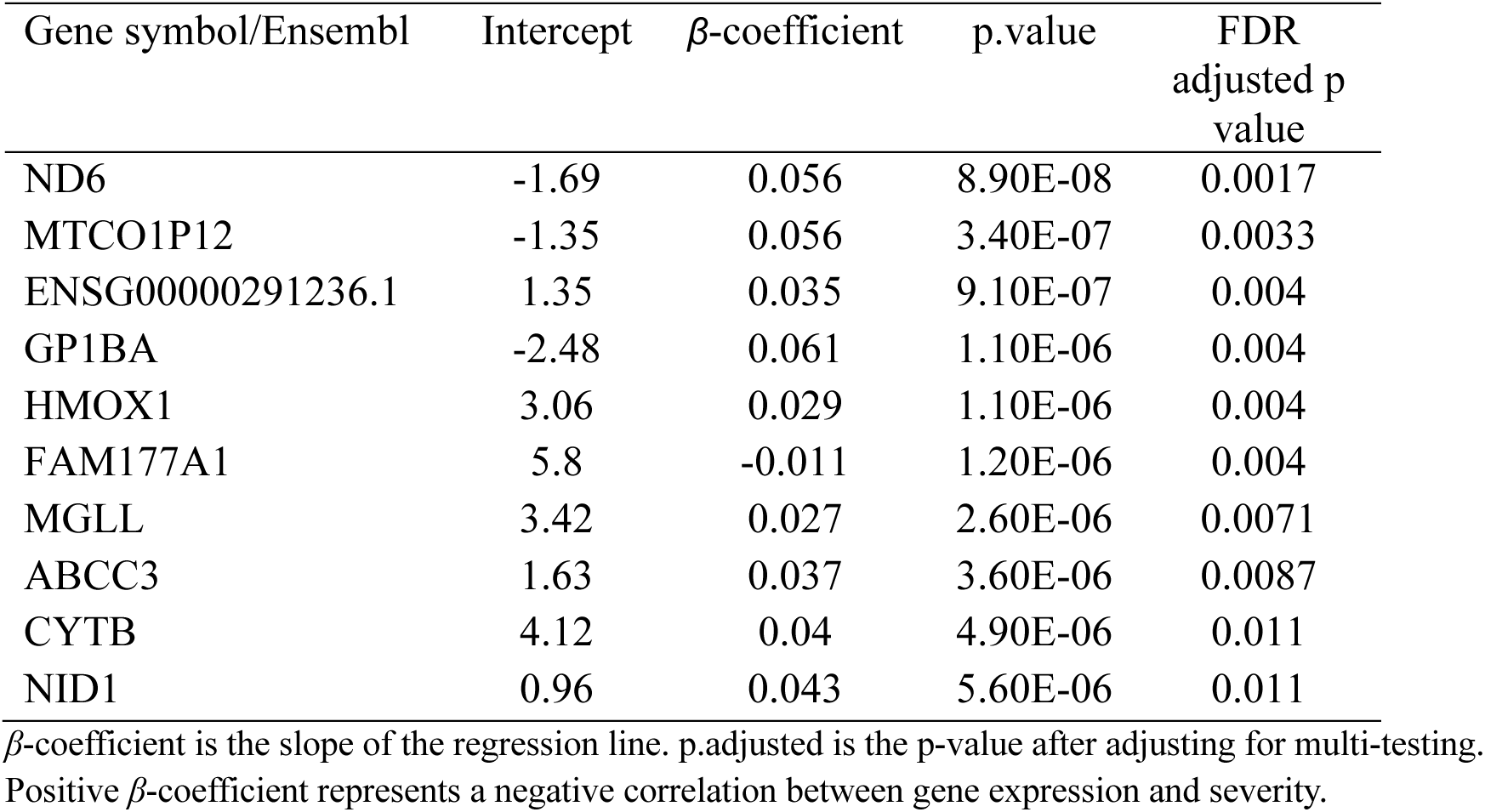
Top 10 representative results (of a total of 144 significant genes) identified in the longitudinal analysis (N_observations_ =103, N_cases_=41).

To evaluate the relevance of these findings tissue specific expression pattern of DE genes were visualized using GTEx (**Supplementary Figure 10**). Tissue expression enrichment (131 recognised out of 144 significant genes) were compared against all tested genes (as the background, 17,274 recognised of 19,260). Downregulated genes were enriched for expression in the brain (cerebellar hemisphere) and the pituitary (Bonferroni-adjusted p < 0.05). In contrast, no upregulated genes were enriched for expression (**Figure 4**).

**Figure 4.**
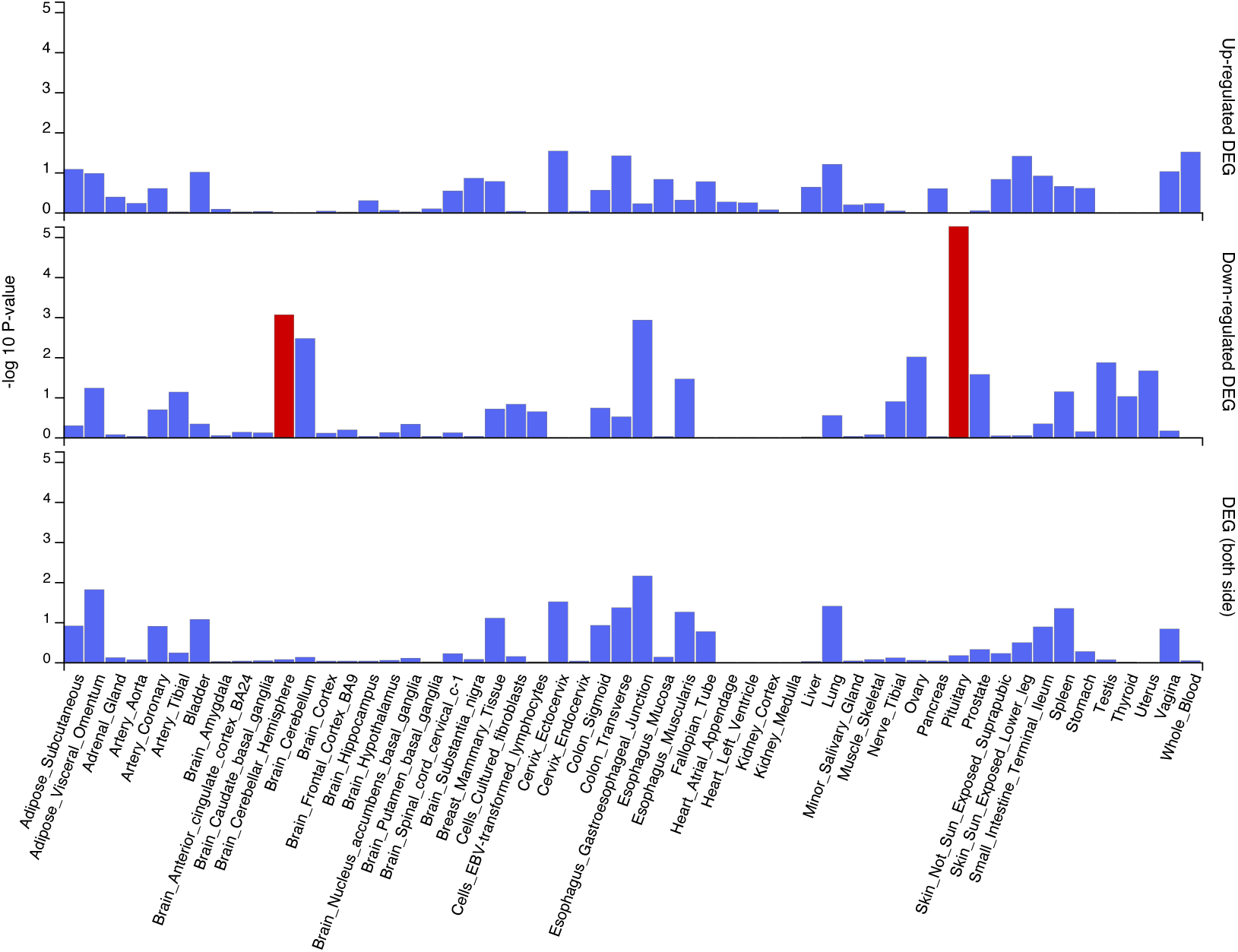
Significantly enriched DEG sets of recognised genes (131/144) in the longitudinal result (compared to all background (17,274 recognised of 19,260)) shows downregulated genes are enriched in brain (cerebellar hemisphere, pituitary) (P_bon_ < 0.05 in red).

Finally, to understand the pathways involved in in longitudinal result, GSEA and Metascape were again used to identify enriched pathways. GSEA identified the 35 enriched pathways, including the p53 pathway (**Figure 3d, Supplementary Table 5**), while Metascape revealed 20 enriched pathways (**Supplementary Figure 11**). Pathways involved in immune response and cell apoptosis appeared in both sets of results.

Using alternative models, we observed few significant findings (FDR of <0.05) (**Supplementary Table 7**). We compared our primary model with model 2, both assessing the association between ALSFRS and gene-expression within-cases, showed highly correlated effect sizes (R^2^=0.71) with reduced p-value inflation in model 2.

### Alternative splicing

The alternative splicing method Leafcutter was used to assess alternative splicing in cases vs. controls. LeafCutter detects novel splicing by clustering junctions without relying on annotations.

A total of 62 clusters were identified in the alternative splicing (AS) analysis (**Supplementary Table 6**) with more than half being cryptic events, i.e., not present in existing annotations. Ten of the identified clusters were in the non-coding DNA regions.

The top hit in AS analyses of cases and controls was a cluster identified in *UBC* (FDR = 1.21e-13, **Figure 5a**). Intersecting all AS results with relevant ALS genes (including risk/similar disease genes), TDP-43 knock-down responsive genes and genes that are loss of function intolerant) included *SETX*, an ALS gene associated with a subtype of ALS [45] (**Figure 5b)**, HLA-A (within the MHC ALS GWAS risk locus) and eight genes that had a high probability of loss of function intolerance score (pLI of ≥ 9) (**Supplementary Table 6**).

**Figure 5.**
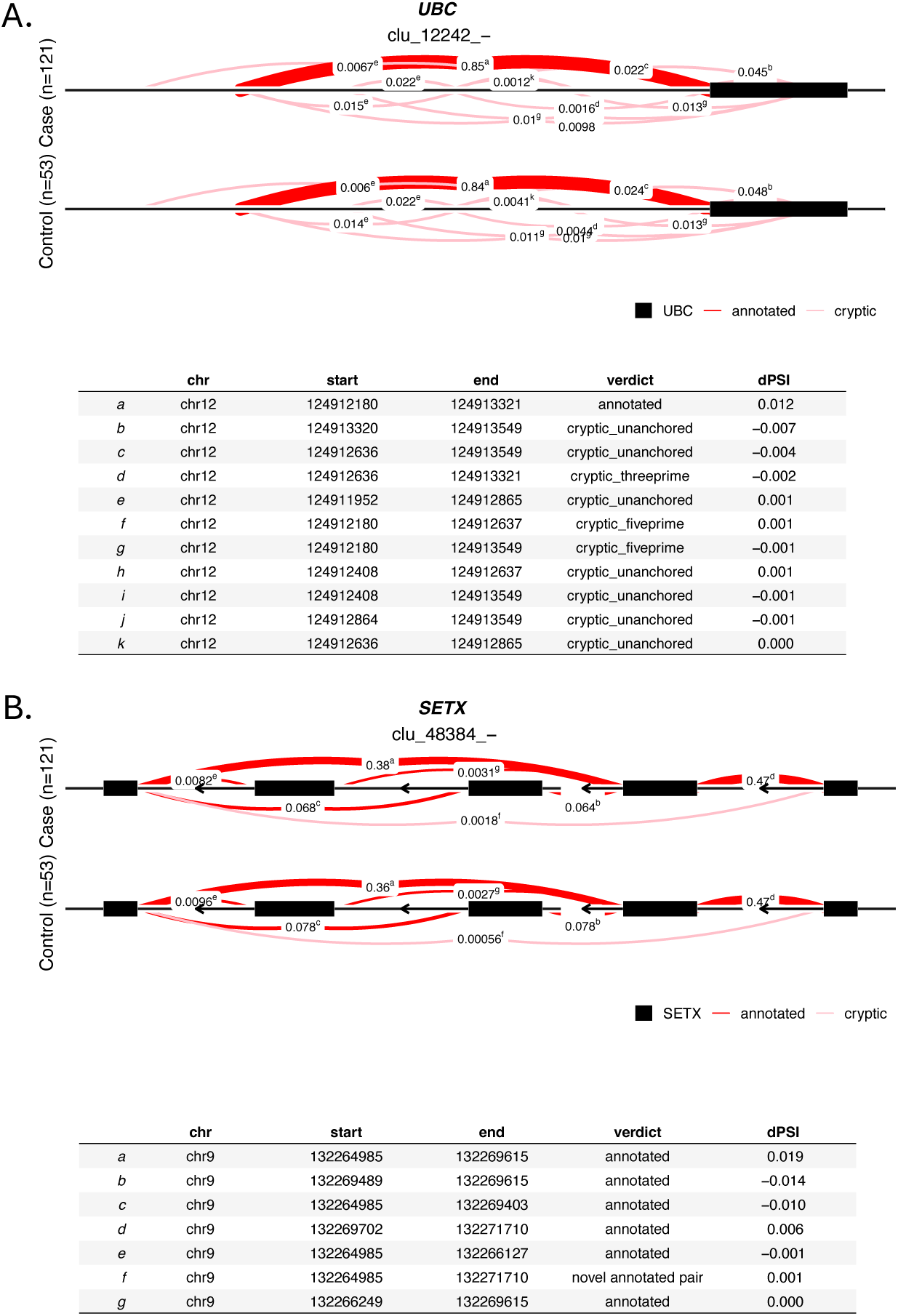
Representative Alternative Splicing Events Detected in ALS Whole Blood using Leafcutter (N_cases_=121 vs N_controls_=53, N=62 significant clusters). A) The top gene from Leafcutter was *UBC* (FDR = 1.21e-13, chr12:124911952-124913549, with 11 introns in the cluster (a-k, coordinates in the table). B) *SETX* (a known Mendelian ALS gene) was the ninth significant AS finding in Leafcutter (FDR = 0.00285, chr9:132264985-132271710, with 7 introns in the cluster, a-g). The mean number of splice junctions supporting each intron is calculated for both conditions and then normalised as a fraction of the total counts (each condition, normalised counts will add up to 1). Each intron is plotted as a line connecting its start and end coordinates with a thickness proportional to the displayed normalised count value. The colour of the intron line indicates whether it is present in the annotation (red) or not (pink). Any exons that are annotated as flanking or being contained within the cluster are added as rectangles to the plot. Clu refers to the intron clusters identified in the study followed by the cluster ID. dPSI: delta percent-spliced-in as calculated by leafcutter.

### Long-read sequencing

A preliminary set of duplicate RNA samples from ALS cases (n=8) and controls (n=8) was also analysed using long-read RNA sequencing (PacBio). The average read length was 1.7kb, with a mean Phred quality score of 38.8 (*SD* ±4.6). The average mapping quality score (MAPQ) across the transcriptome was 59.5 (*SD* ±4.56).

This analysis identified eight significant alternative splicing events, including four intron retentions (from *HLA-DRB1, HLA-DRB5*, and *LST1*; **Supplementary Table 8**) and four exons skipping (from *HLA-DRB5*, *R3HDM4*, *KANSL2;* **Supplementary Table 9**). Differential isoform usage between controls and cases revealed 117 isoforms (46 cryptic) from 87 genes, including 10 HLA family genes (e.g. *HLA-DRB5*, **Figure 6, Supplementary Table 10**)).

**Figure 6.**
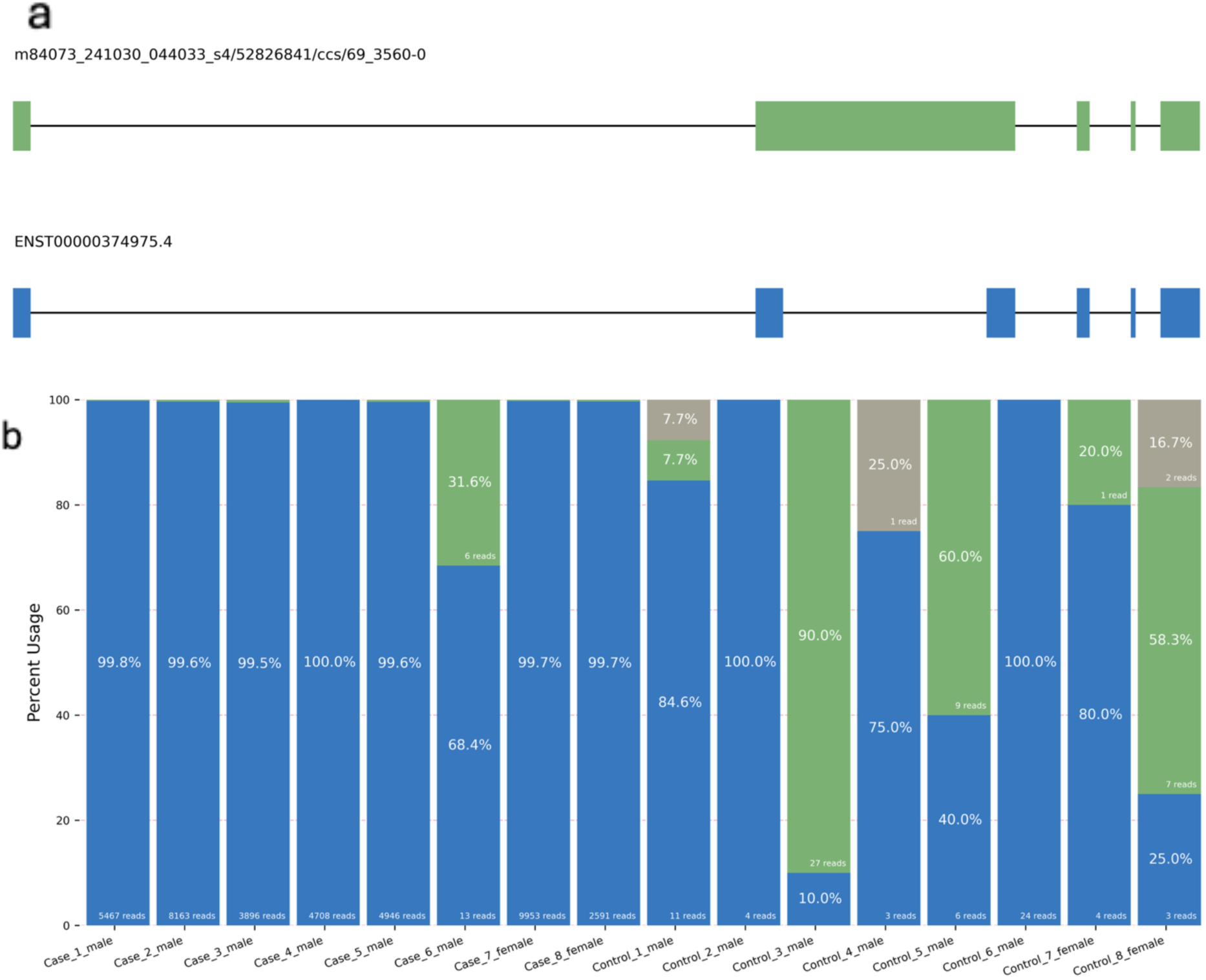
Representative Long-read (PacBio) transcriptome result from N_cases_=8, N_controls_=8. Top hit differentially expressed is *HLA-DRB5* **a)** Visual of the two primary isoforms of *HLA-DRB5* detected in the cohort. **b)** Isoform usage for each control and case (per bar) with respective percent and read counts. Blue area indicates the proportion of isoform ENST00000374975.4, green a cryptic isoform visualised in part a, grey (not shown in a.) another cryptic isoform.

Eight DE genes were also detected, with *HLA-DRB5* showing the strongest signal (log_2_FC > 8; **Supplementary Table 11**). Given the smaller dataset, lower coverage but enhanced resolution of complex regions (i.e. MHC) it was not unexpected that the DE gene detection differed between short-read and long-read results.

### Comparison of cell-type distribution among ALS and control samples

To test for potential cellular heterogeneity in whole blood short-read dataset, we performed cell type deconvolution analysis. Neutrophils represented the largest proportion in both controls and cases (both ∼ 0.48), followed by T cells (at ∼ 0.31 – 0.32) (**Supplementary Figure 12**). Monocytes, B cells, and NK cells made up approximately ∼ 0.19 of the total proportion. However, no statistically significant difference was observed between controls and cases across any of the five cell types (**Supplementary Figure 13**).

### Gene/transcript validation in external blood, iPSC-MN and postmortem RNA-seq data

To validate the results in this study, external publicly available RNA-seq data from large ALS case and control cohorts were utilised. This included blood [40], iPSC-MN [41] and postmortem spinal cord [41].

The blood dataset (N_cases_=96, N_controls_=48) was used for validation [40]. Libraries had been prepared using a different kit (TruSeq Stranded mRNA Sample Prep) and sequencing platform (Illumina HiSeq 2000 platform, 2 × 125bp paired end reads) but data were reprocessed using the same pipeline. Due to lower sequencing depth and technical differences (i.e. library preparation kits) meta-analysis was not deemed relevant (see **Supplementary Note**). A good correlation was still present (*β*-coefficient of the regression line = 0.42 (p-value < 2.2e-16), R^2^ = 0.21) between the datasets indicating shared signals (**Figure 2c; Supplementary Figures 14-15**) but no overlap in significant DE genes (**Supplementary Table 12**).

The largest available iPSC-MN dataset contained a modest number of 43 DE genes [41]. None was found to be common in our DEG list however 55% of the genes had the same direction of effect (more than expected by chance, p-value < 2.2e-16, with a total effect size correlation between the datasets of 0.053). Additionally, the p53 pathway was reported to be enriched in the study [41], which was concordant with our findings (**Figure 2b**).

The largest available post-mortem spinal cord dataset (N = 271), after correction for multiple testing, reported a significant number of 15,874 DE genes due to the extensive neurodegeneration and associated molecular changes characteristic of end-stage ALS [41]. Of these, seven overlapped with our blood DEGs (short read) and showed the same direction of effect. Pathway enrichment found the p53 pathway was upregulated in the spinal cord tissue supporting a shared disease relevant signal [41].

## Discussion

In this study, we present one of the largest blood-based RNA-seq resources in ALS to date, combining both short- and long-read sequencing with an extensive set of analyses to capture both gene expression and transcript-level changes. ALS risk and progression may involve systemic as well as central nervous system pathology and thus blood-based transcriptomics can offers a minimally invasive window into these disease-related biological changes.

Our case-control cohort (N_total_=174) was the largest to date, enhancing sensitivity, the probability of detecting changes between controls and cases, while maintaining stringency. Despite blood not being the primary site of disease, ALS cases showed altered isoforms and differing transcript expression compared to controls. In all, 50 differentially expressed genes were identified, with the most significant change showing specific expression pattern to brain and neurons relevant for further investigation. Other hits in the top 10 include downregulation of *NRG1*, a gene known to be involved in neuron-glia signalling and synaptic plasticity and has been. It plays a role in neuromuscular junction formation and maintenance and has been explored in ALS models as well as several other neurological disorders. *XPO1* was also detected. This is a known nuclear export mediator of proteins and RNA and has been a therapeutic target in neurodegenerative disease models. Thirdly, *AMPH*. This is involved in synaptic vesicle endocytosis, with autoantibodies found in some neurological syndromes and dysfunction of its synaptic trafficking has been implicated in motor neuron degeneration. Overall expression patterns of DE genes however were mixed, demonstrating tissue-wide impacts of disease, with a number of these genes showing same direction of effect in prior smaller / lower coverage blood-transcriptome study [40]. We note that a number of the significant findings were uncharacterised (n=9/50), with function/expression not well reported. While they impose a challenge on understanding how they relate to the pathogenesis of the disease, they also may provide new avenues of understanding novel mechanisms underlying the disease that are not currently known.

Pathway enrichment analyses of the differentially expressed genes revealed changes in the regulation of the immune system and RNA processing in ALS (**Supplementary Table 3, Supplementary Figure 7**). Gene set enrichment analysis (GSEA) identified the top two enriched pathways as the interferon-alpha and interferon-gamma response pathways. This aligns with findings from a PBMC-based transcriptomic study of classic ALS cases (48 sALS, 19 controls; [46]), and with prior molecular investigations linking TDP43 pathology with immune activation [47, 48]. Similar immune-related signatures have been reported in ALS models [47, 49], reinforcing the connection between innate immune response and ALS pathophysiology [48, 49]. Our results also replicate immune- and RNA processing–related pathways reported in a previous blood-based ALS study consistent with the positive correlation in our results [40]. These findings are consistent with the broader literature implicating immune dysregulation [47] and abnormal RNA processing [50, 51] in ALS. Additionally, enrichment of the p53 pathway, involved in cell cycle progression and apoptosis, was observed—supporting previous findings in ALS-affected neurons and tissues [41]. Further investigation in these areas may expand our understanding on the disease.

While immune pathway alterations were detected, there were no significant differences in immune cell proportions using cell-type deconvolution. Given the variability in cell-proportion estimates (**Supplementary Figure 12**), the limitations of reference-based deconvolution methods [52], and prior findings of altered blood composition (i.e. DNA methylation data [53]) we remain cautious about this approach/result.

Our longitudinal case cohort consisting of over 100 observations in N=41 ALS cases was the largest blood RNAseq progression cohort to date. We identified close to 150 genes associated with disease progression in this cohort (**Table 3**). A majority showed a decline in expression along with disease progression. Interestingly, there was no overlap between the longitudinal and case-control analyses. This aligns with prior genetic studies showing that the mechanisms underlying disease risk and disease progression may be distinct, pointing to independent biological pathways [4]. For the pathway analyses, cell proliferation pathways were enriched. Consensus in literature on how the pathways are involved and whether in general proliferation rates are increased or reduced in ALS has not been reached [54, 55]. Downregulated genes detected in blood were significantly enriched in expression in the cerebellar hemisphere, a brain region relevant to motor control, coordination, and emotional/motivational processing. Additionally, there was enrichment in non-brain tissue, the pituitary which is a tissue involved in hormonal regulation, metabolism, and immune function. These peripheral blood signals appear sensitive to both CNS and non-CNS relevant transcriptional changes.

We note that some inflation was observed in the longitudinal analyses, which is not uncommon in repeated measures data. However, variance partitioning revealed that much variance remained unexplained by the model in the dataset (**Supplementary Figure 9**). One possible explanation is the limited resolution of ALSFRS-R in capturing physical status, complementing this with alternative metrics such as Neurofilament light chain (NfL) concentration [56] or actigraphy [57] may improve the robustness and replicability of future analyses. Despite the modest cohort size-these longitudinal datasets are essential to build for disease trajectories and capturing individual heterogeneity in ALS progression.

To address growing evidence that RNA dysregulation is a key contributor to ALS pathogenesis, we leveraged deeply sequenced short-read data to examine alternative splicing using a widely adopted tool LeafCutter, which can detect unannotated, local splicing variation. This identified 62 significantly altered splicing clusters (**Table 4**, **Supplementary Table 6**). Notably, *UNC13A* or *STMN2* were not detected in our splice results with low/absent expression in our blood data (not unexpected based on external data [42]). Interestingly, our top splicing signal was in *UBC*, which encodes a which encodes ubiquitin C—a key component of the ubiquitin-proteasome system involved in protein degradation, cellular stress responses, and regulation of immune signalling. Alternative splicing of UBC may reflect broader dysregulation of proteostasis and inflammatory pathways in ALS

**Table 4.** Top 10 representative results of 62 significantly identified ALS clusters in the alternative splicing analysis.

| Gene symbol | Coordinate | Annotation | FDR | pLI ≥ 0.9 | ALS related |
| --- | --- | --- | --- | --- | --- |
| UBC | chr12:124911952-124913549 | cryptic | 1.21E-13 |  |  |
| RSRP1 | chr1:25243633-25245150 | cryptic | 0.0000357 |  |  |
| . | chr21:8212485-8439743 | annotated | 0.0000798 |  |  |
| HLA-A | chr6:29828818-30491137 | cryptic | 0.000138 |  | Y |
| . | chr1:496286-737774 | annotated | 0.000138 |  |  |
| ENSG00000290937 | chr3:198153409-198180755 | cryptic | 0.000343 |  |  |
| . | chr6:29850126-30007576 | annotated | 0.000513 |  |  |
| NSFL1C | chr20:1458274-1479994 | cryptic | 0.00116 | Y |  |
| SETX | chr9:132264985-132271710 | cryptic | 0.00285 | Y | Y |
| EWSR1 | chr22:29268349-29272380 | cryptic | 0.00648 | Y | Y |

The dots (.) in the gene symbol column are non-coding DNA regions. pLI is the probability of loss of function intolerance score. Y=yes.

This study also presents a preliminary blood-based long-read RNA-seq dataset (the first that we know of) offering a valuable preliminary look at transcriptomic complexity in this peripheral tissue. While fewer differential expression and splicing events were detected compared to short-read data— likely due to limited sample size 16 —the long-read seq did effectively resolve complex genomic regions, including multiple splicing events in *HLA-DRB1* and *HLA-DRB5*. Notably, *HLA-DRB5* showed a strong case–control expression difference, consistent with prior genetic associations in ALS [58]. As long-read datasets grow, their potential to reveal novel disease-relevant mechanisms will increase.

Importantly, our study results contain direction of effect. This can be utilised for functional follow-up with a number of drub-bank candidates that could be relevant (**Supplementary Tables 13-14**).

Our study has a number of limitations. The transcriptome data derived from whole blood can reflect systemic physiology given it is not the primary pathology site. Moreover, although blood is a clinically accessible and practical biospecimen, technical and biological variability, including sample collection, processing time, and medication or comorbidities, may contribute to noise in gene expression data.

Despite these limitations, our study revealed biologically meaningful findings suggestive of disease-relevant processes. Notably, enrichment of the p53 pathway (**Figure 2b**) was consistent with signals detected in the largest meta-analysis of ALS cells and post-mortem CNS tissue to date [41] and the innate immune system [48, 49]. This supports the possibility of capturing central disease processes via peripheral blood profiling, particularly if these pathways become therapeutic targets or biomarkers of treatment response. This includes direction of effect which can be utilised for functional follow-up with a number of drug-bank candidates that could be relevant (**Supplementary Tables 13-14**). We also identified alternative splicing events, including in the ALS-gene *SETX* (associated with Mendelian inheritance), noting the magnitude of splicing changes was subtle, reinforcing the need for follow-up, including in disease-relevant cells/tissues.

Our longitudinal and long-read sequencing datasets provided additional insight into transcriptomic changes over time in CNS and non-CNS tissues and splicing complexity in ALS. While these analyses were limited by modest sample sizes and variable follow-up intervals, they provide a basis for future studies with appropriately large, harmonized cohorts to validate and expand on these observations.

Blood-based transcriptomics can offer a minimally invasive window into disease-related biological processes. The findings we present show that ALS risk and progression may involve systemic, as well as central nervous system, pathology. This comprehensive dataset enables exploration of the peripheral transcriptome, capturing immune and systemic responses to neurodegeneration—including previously recognised RNA dysregulation—and reflects the known heterogeneity of ALS. Importantly, we identified numerous novel findings, including differential usage of previously unannotated transcripts and splicing events, several of which implicate genes and pathways relevant to ALS biology. These discoveries warrant further functional validation in disease-relevant tissues and models. Together, our work demonstrates the value of integrative RNA sequencing approaches in blood to further dissect the molecular mechanisms of ALS.

## Funding/Acknowledgements

We gratefully acknowledge all participants and their families for generously contributing to this research. We also thank the clinical teams and staff involved in supporting sample and data collection who made this project possible. The data generated in this project was funded by an IMPACT grant from FightMND (2022, to FCG). Additional project funding and support were provided by a Daniel McLoone MND Research Grant from Motor Neurone Disease Australia (IG2312, to FCG), (Australian) National Health and Medical Research Council (grants 1078901, 1113400, 1087889) to NRW and the MND Australia Ice Bucket Challenge Grant (to NRW). FCG was funded by a Scott Sullivan Fellowship (MND and Me/MNDRA). We thank Kelly Williams and Natalie Grima for their helpful responses re: GSE234297.

## Data availability

The external RNA-seq datasets used for validation were publicly available. The blood RNAseq dataset was downloaded from NCBI GEO (accession GSE234297, [40]) and the results from iPSC-MN and postmortem tissue RNAseq analysis was taken from Ziff et al. [41]. The RNA-seq blood data generated here is available from the author upon reasonable request; an online submission for upload is in progress.

## References

1. Kiernan MC, Vucic S, Cheah BC, Turner MR, Eisen A, Hardiman O, Burrell JR, Zoing MC: Amyotrophic lateral sclerosis. Lancet (London, England) 2011, 377:942–955.

2. Masrori P, Van Damme P: Amyotrophic lateral sclerosis: a clinical review. Eur J Neurol 2020, 27:1918–1929.

3. Nijs M, Van Damme P: The genetics of amyotrophic lateral sclerosis. Curr Opin Neurol 2024, 37:560–569.

4. van Rheenen W, van der Spek RAA, Bakker MK, van Vugt J, Hop PJ, Zwamborn RAJ, de Klein N, Westra HJ, Bakker OB, Deelen P, et al: Common and rare variant association analyses in amyotrophic lateral sclerosis identify 15 risk loci with distinct genetic architectures and neuron-specific biology. Nat Genet 2021, 53:1636–1648.

5. Restuadi R, Steyn FJ, Kabashi E, Ngo ST, Cheng FF, Nabais MF, Thompson MJ, Qi T, Wu Y, Henders AK, et al: Functional characterisation of the amyotrophic lateral sclerosis risk locus GPX3/TNIP1. Genome Med 2022, 14:7.

6. Taylor JP, Brown RH, Jr., Cleveland DW: Decoding ALS: from genes to mechanism. Nature 2016, 539:197–206.

7. Brown AL, Wilkins OG, Keuss MJ, Hill SE, Zanovello M, Lee WC, Bampton A, Lee FCY, Masino L, Qi YA, et al: Author Correction: TDP-43 loss and ALS-risk SNPs drive mis-splicing and depletion of UNC13A. Nature 2024, 631:E7.

8. Ma XR, Prudencio M, Koike Y, Vatsavayai SC, Kim G, Harbinski F, Briner A, Rodriguez CM, Guo C, Akiyama T, et al: TDP-43 represses cryptic exon inclusion in the FTD-ALS gene UNC13A. Nature 2022, 603:124–130.

9. Elden AC, Kim HJ, Hart MP, Chen-Plotkin AS, Johnson BS, Fang X, Armakola M, Geser F, Greene R, Lu MM, et al: Ataxin-2 intermediate-length polyglutamine expansions are associated with increased risk for ALS. Nature 2010, 466:1069–1075.

10. Barker HV, Niblock M, Lee YB, Shaw CE, Gallo JM: RNA Misprocessing in C9orf72-Linked Neurodegeneration. Front Cell Neurosci 2017, 11:195.

11. Sreedharan J, Blair IP, Tripathi VB, Hu X, Vance C, Rogelj B, Ackerley S, Durnall JC, Williams KL, Buratti E, et al: TDP-43 mutations in familial and sporadic amyotrophic lateral sclerosis. Science 2008, 319:1668–1672.

12. Kabashi E, Valdmanis PN, Dion P, Spiegelman D, McConkey BJ, Vande Velde C, Bouchard JP, Lacomblez L, Pochigaeva K, Salachas F, et al: TARDBP mutations in individuals with sporadic and familial amyotrophic lateral sclerosis. Nat Genet 2008, 40:572–574.

13. Vance C, Rogelj B, Hortobagyi T, De Vos KJ, Nishimura AL, Sreedharan J, Hu X, Smith B, Ruddy D, Wright P, et al: Mutations in FUS, an RNA processing protein, cause familial amyotrophic lateral sclerosis type 6. Science 2009, 323:1208–1211.

14. Mann JR, Gleixner AM, Mauna JC, Gomes E, DeChellis-Marks MR, Needham PG, Copley KE, Hurtle B, Portz B, Pyles NJ, et al: RNA Binding Antagonizes Neurotoxic Phase Transitions of TDP-43. Neuron 2019, 102:321–338 e328.

15. Lagier-Tourenne C, Polymenidou M, Cleveland DW: TDP-43 and FUS/TLS: emerging roles in RNA processing and neurodegeneration. Hum Mol Genet 2010, 19:R46–64.

16. Stark R, Grzelak M, Hadfield J: RNA sequencing: the teenage years. Nat Rev Genet 2019, 20:631–656.

17. Cai G, Li H, Lu Y, Huang X, Lee J, Muller P, Ji Y, Liang S: Accuracy of RNA-Seq and its dependence on sequencing depth. BMC Bioinformatics 2012, 13 Suppl 13:S5.

18. Liu Y, Zhou J, White KP: RNA-seq differential expression studies: more sequence or more replication? Bioinformatics 2014, 30:301–304.

19. Fresard L, Smail C, Ferraro NM, Teran NA, Li X, Smith KS, Bonner D, Kernohan KD, Marwaha S, Zappala Z, et al: Identification of rare-disease genes using blood transcriptome sequencing and large control cohorts. Nat Med 2019, 25:911–919.

20. Chen S, Zhou Y, Chen Y, Gu J: fastp: an ultra-fast all-in-one FASTQ preprocessor. Bioinformatics 2018, 34:i884–i890.

21. Dobin A, Davis CA, Schlesinger F, Drenkow J, Zaleski C, Jha S, Batut P, Chaisson M, Gingeras TR: STAR: ultrafast universal RNA-seq aligner. Bioinformatics 2013, 29:15–21.

22. Liao Y, Smyth GK, Shi W: featureCounts: an efficient general purpose program for assigning sequence reads to genomic features. Bioinformatics 2014, 30:923–930.

23. Love MI, Huber W, Anders S: Moderated estimation of fold change and dispersion for RNA-seq data with DESeq2. Genome Biol 2014, 15:550.

24. Gene Ontology C, Aleksander SA, Balhoff J, Carbon S, Cherry JM, Drabkin HJ, Ebert D, Feuermann M, Gaudet P, Harris NL, et al: The Gene Ontology knowledgebase in 2023. Genetics 2023, 224.

25. Kanehisa M, Furumichi M, Sato Y, Matsuura Y, Ishiguro-Watanabe M: KEGG: biological systems database as a model of the real world. Nucleic Acids Res 2025, 53:D672–D677.

26. Milacic M, Beavers D, Conley P, Gong C, Gillespie M, Griss J, Haw R, Jassal B, Matthews L, May B, et al: The Reactome Pathway Knowledgebase 2024. Nucleic Acids Res 2024, 52:D672–D678.

27. Ruepp A, Waegele B, Lechner M, Brauner B, Dunger-Kaltenbach I, Fobo G, Frishman G, Montrone C, Mewes HW: CORUM: the comprehensive resource of mammalian protein complexes--2009. Nucleic Acids Res 2010, 38:D497–501.

28. Dolgalev I: msigdbr: MSigDB gene sets for multiple organisms in a tidy data format. R package version 2022, 7.

29. Sergushichev AA: An algorithm for fast preranked gene set enrichment analysis using cumulative statistic calculation. bioRxiv 2016:060012.

30. Subramanian A, Tamayo P, Mootha VK, Mukherjee S, Ebert BL, Gillette MA, Paulovich A, Pomeroy SL, Golub TR, Lander ES, Mesirov JP: Gene set enrichment analysis: a knowledge-based approach for interpreting genome-wide expression profiles. Proc Natl Acad Sci U S A 2005, 102:15545–15550.

31. Hoffman GE, Schadt EE: variancePartition: interpreting drivers of variation in complex gene expression studies. BMC Bioinformatics 2016, 17:483.

32. Li YI, Knowles DA, Humphrey J, Barbeira AN, Dickinson SP, Im HK, Pritchard JK: Annotation-free quantification of RNA splicing using LeafCutter. Nat Genet 2018, 50:151–158.

33. Karczewski KJ, Francioli LC, Tiao G, Cummings BB, Alfoldi J, Wang Q, Collins RL, Laricchia KM, Ganna A, Birnbaum DP, et al: The mutational constraint spectrum quantified from variation in 141,456 humans. Nature 2020, 581:434–443.

34. Klim JR, Williams LA, Limone F, Guerra San Juan I, Davis-Dusenbery BN, Mordes DA, Burberry A, Steinbaugh MJ, Gamage KK, Kirchner R, et al: ALS-implicated protein TDP-43 sustains levels of STMN2, a mediator of motor neuron growth and repair. Nat Neurosci 2019, 22:167–179.

35. Garton FC, Benyamin B, Zhao Q, Liu Z, Gratten J, Henders AK, Zhang ZH, Edson J, Furlong S, Morgan S, et al: Whole exome sequencing and DNA methylation analysis in a clinical amyotrophic lateral sclerosis cohort. Mol Genet Genomic Med 2017, 5:418–428.

36. Tang AD, Soulette CM, van Baren MJ, Hart K, Hrabeta-Robinson E, Wu CJ, Brooks AN: Full-length transcript characterization of SF3B1 mutation in chronic lymphocytic leukemia reveals downregulation of retained introns. Nat Commun 2020, 11:1438.

37. Newman AM, Steen CB, Liu CL, Gentles AJ, Chaudhuri AA, Scherer F, Khodadoust MS, Esfahani MS, Luca BA, Steiner D, et al: Determining cell type abundance and expression from bulk tissues with digital cytometry. Nat Biotechnol 2019, 37:773–782.

38. Patro R, Duggal G, Love MI, Irizarry RA, Kingsford C: Salmon provides fast and bias-aware quantification of transcript expression. Nat Methods 2017, 14:417–419.

39. Soneson C, Love MI, Robinson MD: Differential analyses for RNA-seq: transcript-level estimates improve gene-level inferences. F1000Res 2015, 4:1521.

40. Grima N, Liu S, Southwood D, Henden L, Smith A, Lee A, Rowe DB, D’Silva S, Blair IP, Williams KL: RNA sequencing of peripheral blood in amyotrophic lateral sclerosis reveals distinct molecular subtypes: Considerations for biomarker discovery. Neuropathol Appl Neurobiol 2023, 49:e12943.

41. Ziff OJ, Neeves J, Mitchell J, Tyzack G, Martinez-Ruiz C, Luisier R, Chakrabarti AM, McGranahan N, Litchfield K, Boulton SJ, et al: Integrated transcriptome landscape of ALS identifies genome instability linked to TDP-43 pathology. Nat Commun 2023, 14:2176.

42. Consortium GT: The Genotype-Tissue Expression (GTEx) project. Nat Genet 2013, 45:580–585.

43. Uhlen M, Fagerberg L, Hallstrom BM, Lindskog C, Oksvold P, Mardinoglu A, Sivertsson A, Kampf C, Sjostedt E, Asplund A, et al: Proteomics. Tissue-based map of the human proteome. Science 2015, 347:1260419.

44. Liu C, Cripe TP, Kim MO: Statistical issues in longitudinal data analysis for treatment efficacy studies in the biomedical sciences. Mol Ther 2010, 18:1724–1730.

45. Ma L, Shi Y, Chen Z, Li S, Zhang J: A novel SETX gene mutation associated with Juvenile amyotrophic lateral sclerosis. Brain Behav 2018, 8:e01066.

46. Dragoni F, Garofalo M, Di Gerlando R, Rizzo B, Bordoni M, Scarian E, Viola C, Bettoni V, Fiamingo G, Tornabene D, et al: Whole transcriptome analysis of unmutated sporadic ALS patients’ peripheral blood reveals phenotype-specific gene expression signature. Neurobiol Dis 2025, 206:106823.

47. Piccoli T, Castro F, La Bella V, Meraviglia S, Di Simone M, Salemi G, Dieli F, Spataro R: Role of the immune system in amyotrophic lateral sclerosis. Analysis of the natural killer cells and other circulating lymphocytes in a cohort of ALS patients. BMC Neurol 2023, 23:222.

48. Evangelista BA, Ragusa JV, Pellegrino K, Wu Y, Quiroga-Barber IY, Cahalan SR, Arooji OK, Madren JA, Schroeter S, Cozzarin J, et al: TDP-43 pathology links innate and adaptive immunity in amyotrophic lateral sclerosis. bioRxiv 2024.

49. Mimic S, Aru B, Pehlivanoglu C, Sleiman H, Andjus PR, Yanikkaya Demirel G: Immunology of amyotrophic lateral sclerosis - role of the innate and adaptive immunity. Front Neurosci 2023, 17:1277399.

50. Strong MJ: The evidence for altered RNA metabolism in amyotrophic lateral sclerosis (ALS). J Neurol Sci 2010, 288:1–12.

51. Honda N, Watanabe Y, Honda H, Uemoto M, Fukuhara H, Hanajima R: Implications of Mutant SOD1 on RNA Processing and Interferon Responses in Amyotrophic Lateral Sclerosis: Omics Data Analysis. Cureus 2025, 17:e81045.

52. Im Y, Kim Y: A Comprehensive Overview of RNA Deconvolution Methods and Their Application. Mol Cells 2023, 46:99–105.

53. Nabais MF, Lin T, Benyamin B, Williams KL, Garton FC, Vinkhuyzen AAE, Zhang F, Vallerga CL, Restuadi R, Freydenzon A, et al: Significant out-of-sample classification from methylation profile scoring for amyotrophic lateral sclerosis. NPJ Genom Med 2020, 5:10.

54. Riancho J, Castanedo-Vazquez D, Gil-Bea F, Tapia O, Arozamena J, Duran-Vian C, Sedano MJ, Berciano MT, Lopez de Munain A, Lafarga M: ALS-derived fibroblasts exhibit reduced proliferation rate, cytoplasmic TDP-43 aggregation and a higher susceptibility to DNA damage. J Neurol 2020, 267:1291–1299.

55. Chi L, Ke Y, Luo C, Li B, Gozal D, Kalyanaraman B, Liu R: Motor neuron degeneration promotes neural progenitor cell proliferation, migration, and neurogenesis in the spinal cords of amyotrophic lateral sclerosis mice. Stem Cells 2006, 24:34–43.

56. Su WM, Cheng YF, Jiang Z, Duan QQ, Yang TM, Shang HF, Chen YP: Predictors of survival in patients with amyotrophic lateral sclerosis: A large meta-analysis. EBioMedicine 2021, 74:103732.

57. Straczkiewicz M, Burke KM, Calcagno N, Premasiri A, Vieira FG, Onnela JP, Berry JD: Free-living monitoring of ALS progression in upper limbs using wearable accelerometers. J Neuroeng Rehabil 2024, 21:223.

58. Yang X, Zheng J, Tian S, Chen Y, An R, Zhao Q, Xu Y: HLA-DRA/HLA-DRB5 polymorphism affects risk of sporadic ALS and survival in a southwest Chinese cohort. J Neurol Sci 2017, 373:124–128.

