## Supplementary Figures and Note for "A Blood Transcriptomic Resource for ALS Highlights Disease-Associated Signatures and Alternative Splicing Events"

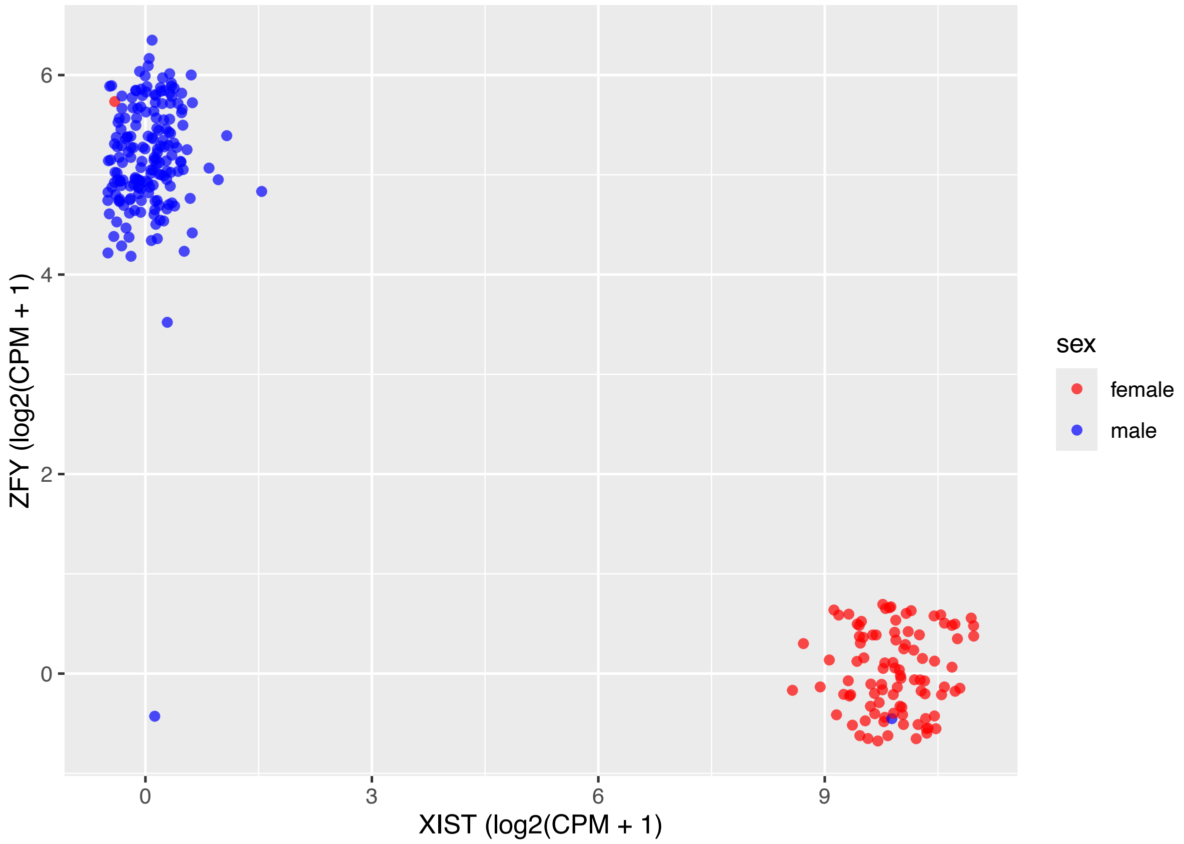


**Supplementary Figure 1**. Expression of two sex related genes identified two sex-mismatched samples in the dataset. Each dot represents a sample and is coloured based on the recorded sex.

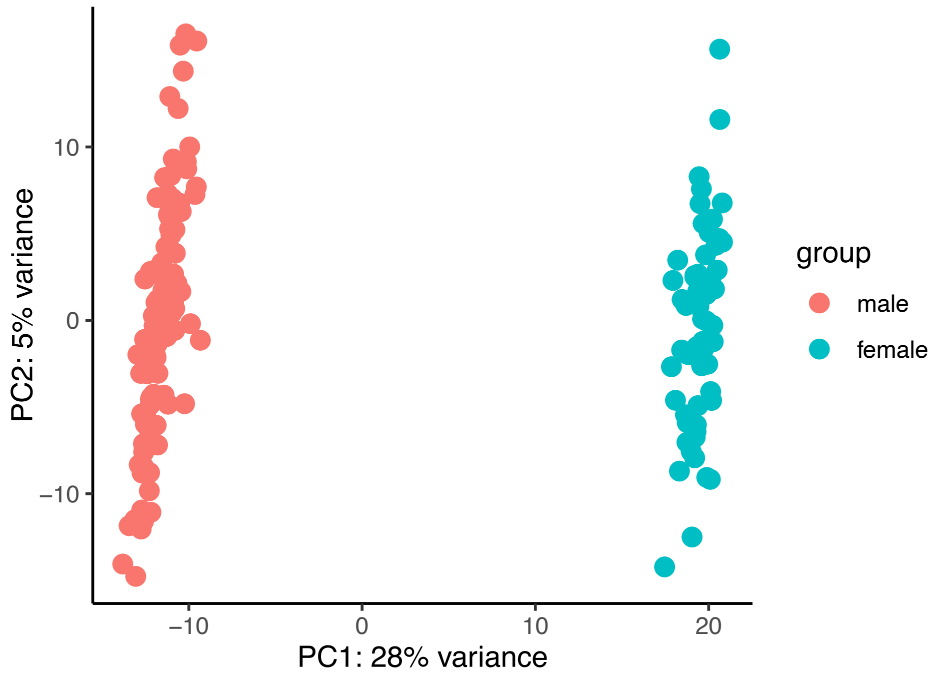


**Supplementary Figure 2.** Principal components (PCA) of the 174 samples used in the differential gene expression analysis after vst normalization (*DEseq2*). Each dot represents a sample and coloured based on clinically recorded sex. Sub-groups based on sex were observed.

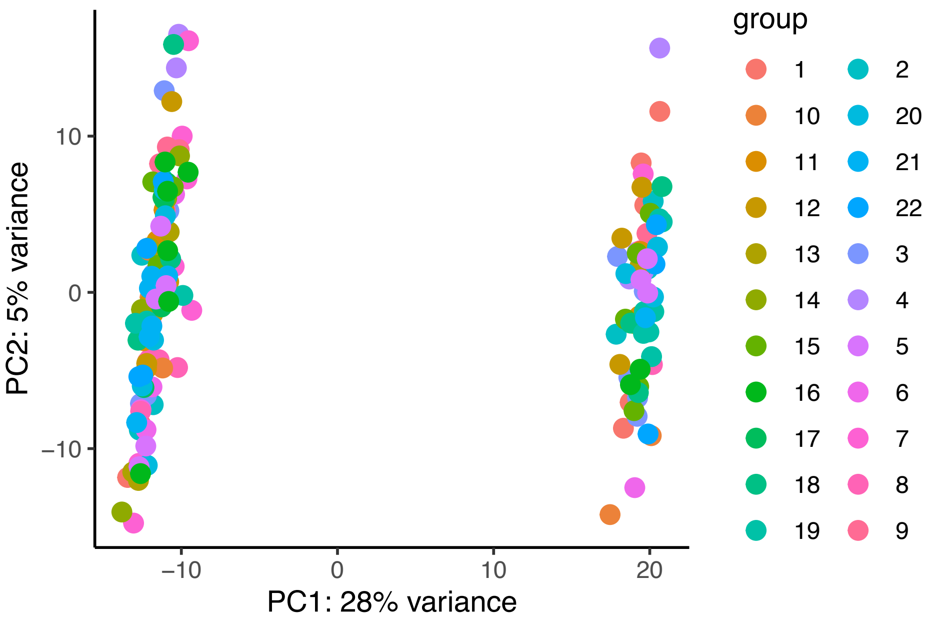


**Supplementary Figure 3**. PCA plot of the 174 samples used in the differential gene expression analysis after vst normalization (*DEseq2*). Each dot represents a sample and coloured based on processing batch. Sub-groups based on batch were not observed.

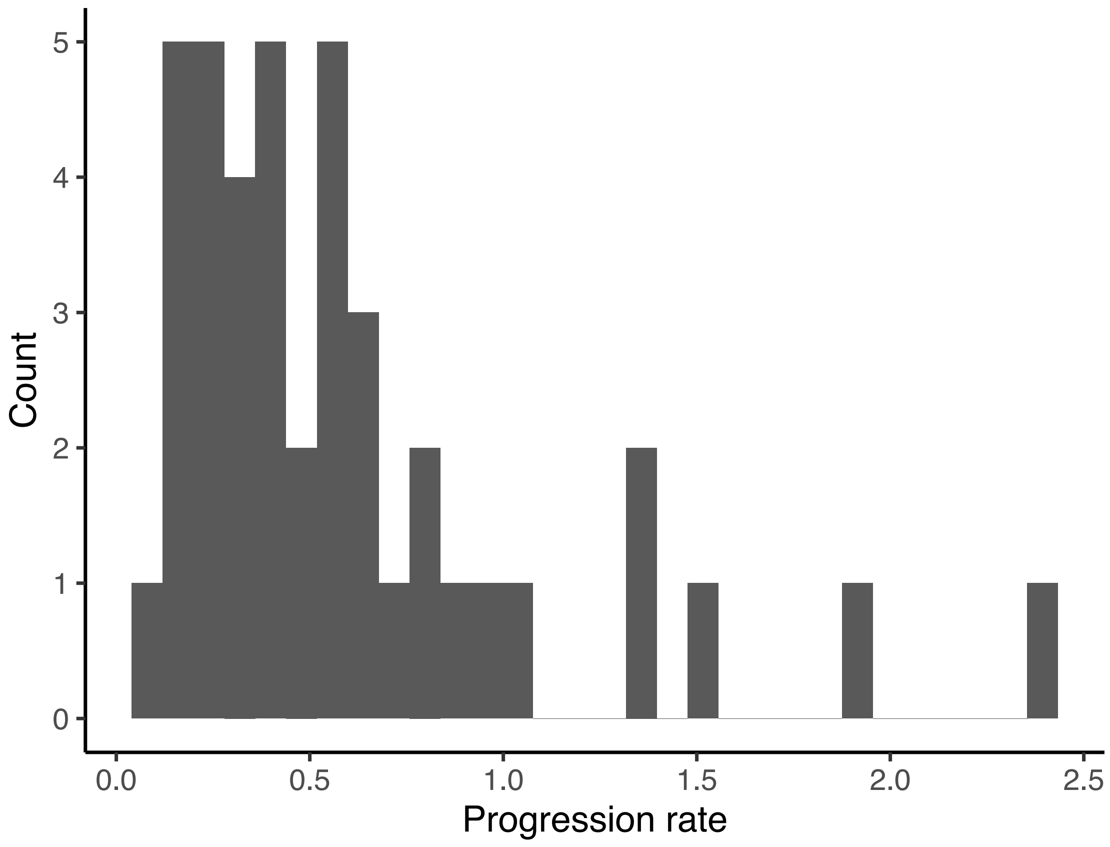


**Supplementary Figure 4.** The average ALS progression rates in the longitudinal cohort. Rates were obtained by finding the difference between onset (ALSFRS-R (48)) and the ALSFRS-R at the last visit of the individual, divided by time (months between onset and last visit).

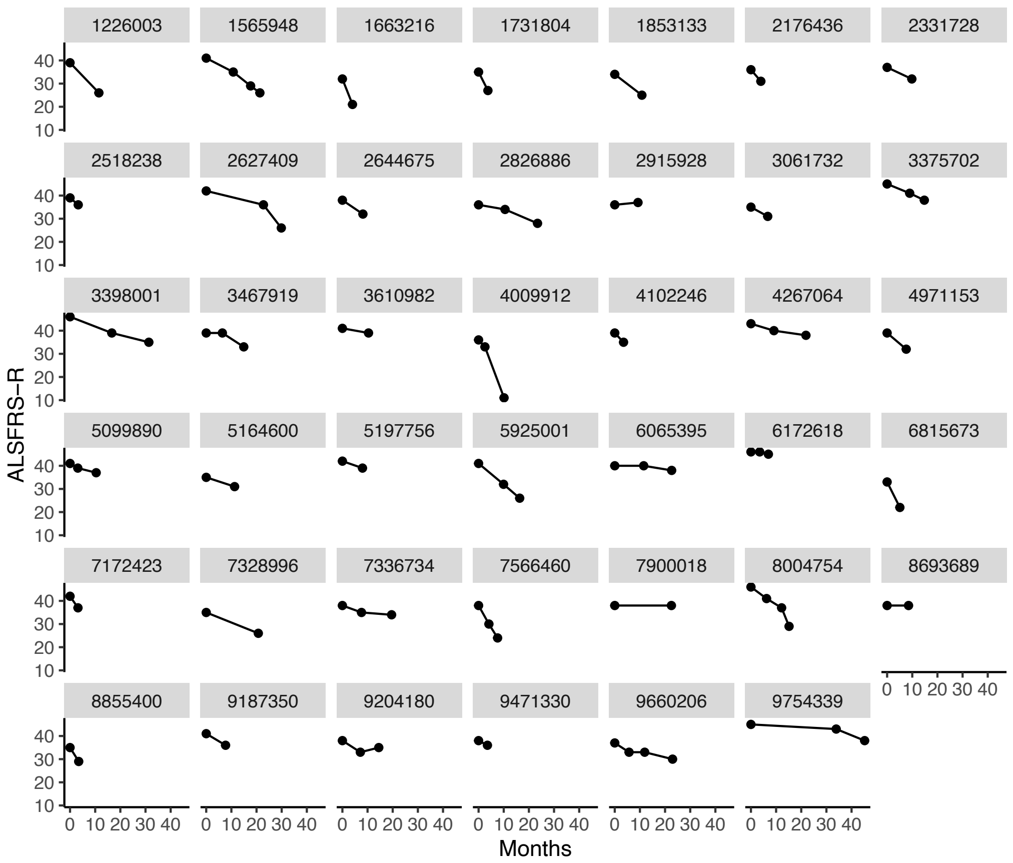


**Supplementary Figure 5.** A panel showing the changes in ALSFRS-R scores of each visit in months. The numbers in the grey boxes are encrypted patient IDs. Each dot represents a visit.

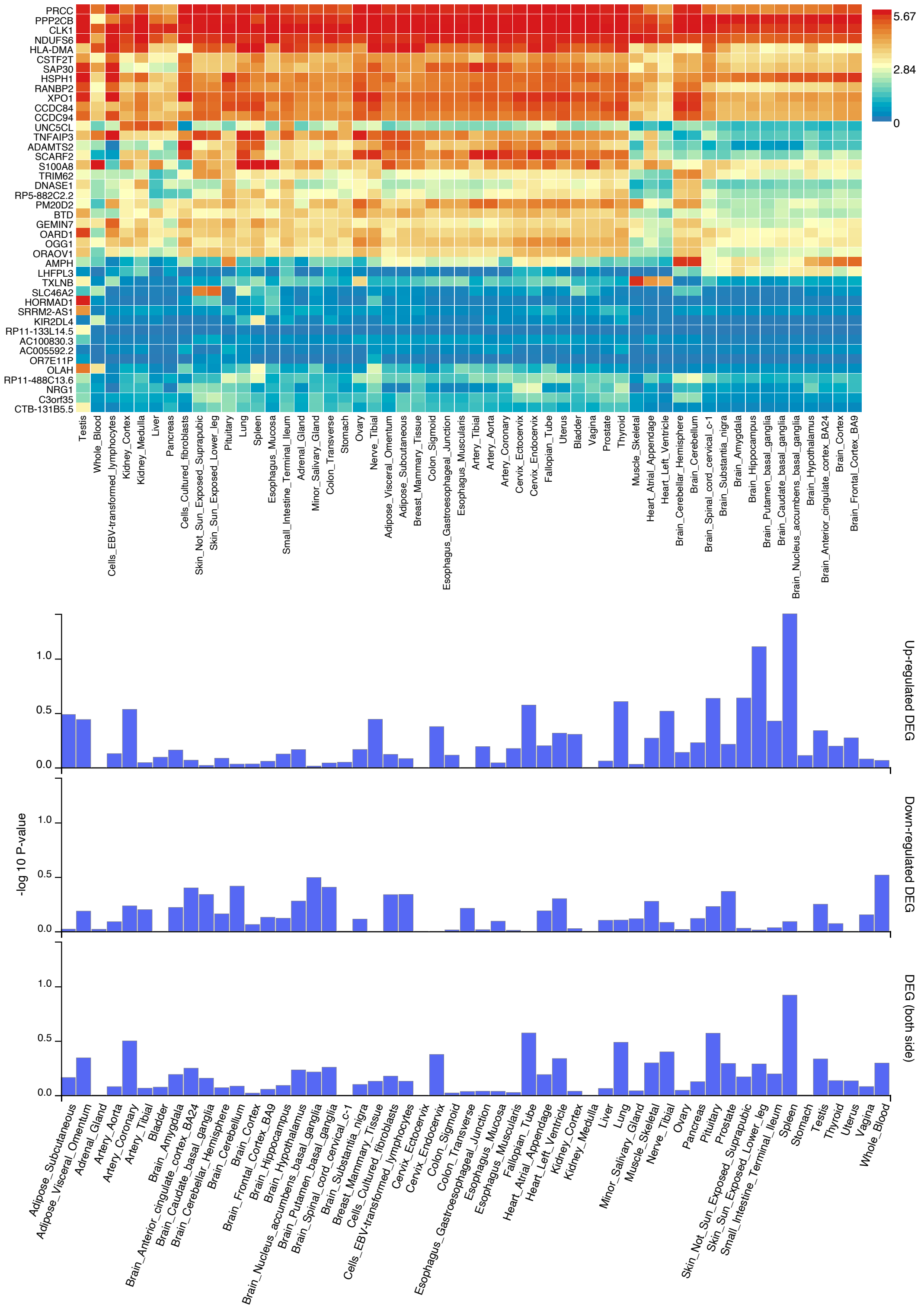


**Supplementary Figure 6**. Differentially expressed (DE) genes do not show a distinct expression pattern across tissues. The heatmap illustrates the correlations of expression patterns of the DE genes across human (GTEx) tissues clustered by tissue and pattern. Colour shade indicates average $\log_2$ expression level of that gene in corresponding tissue (see legend). The bar graphs show no enrichment of up or downregulated genes. Plotted is tissue (x-axis) and the $\log_{10}$ p-value of enrichment of significant genes (45 recognised) compared to all genes in the DE analysis (17,483 recognised).

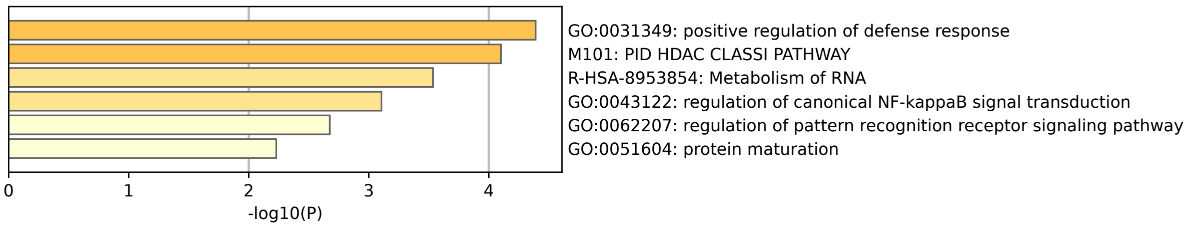


**Supplementary Figure 7**. Result of the pathway enrichment analysis on DE genes using Metascape. A total of 6 pathways were enriched in the DE genes identified in this study.

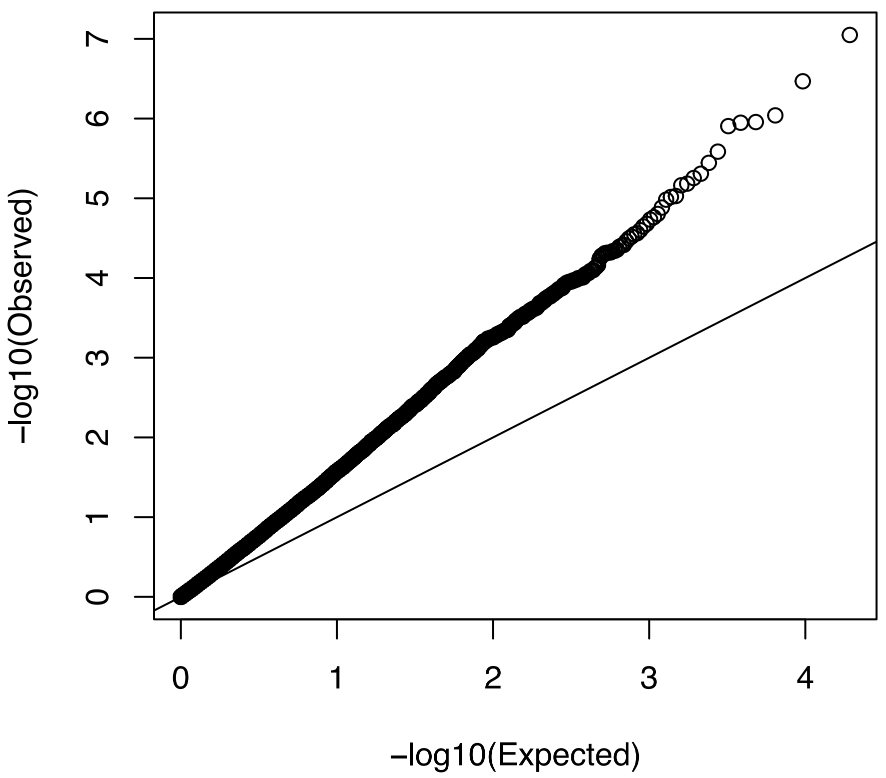


**Supplementary Figure 8.** A quantile–quantile plot using the -log10 transformed p-values of the longitudinal result (progression of disease vs gene expression) compared against a uniform distribution. The diagonal line represents x = y. The inflation factor (λ) is 1.91.

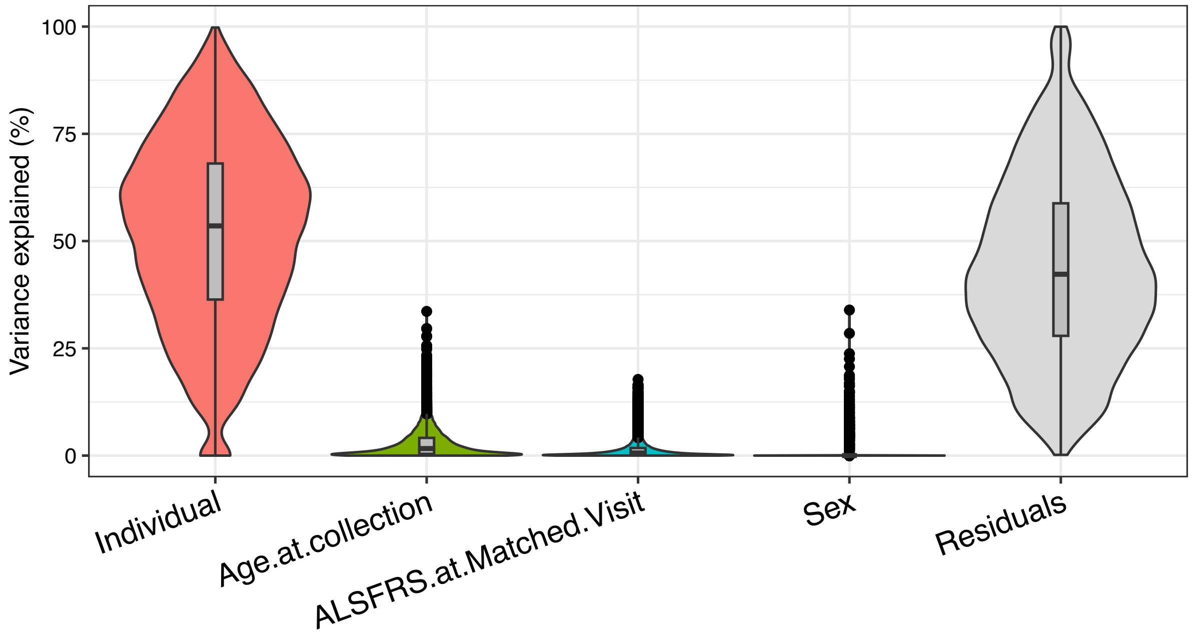


**Supplementary Figure 9**. Variance partitioning analysis on the longitudinal dataset (sample = 103, individuals = 41). Age.at collection is age, ALSFRS.at.Matched.Visit is ALSFRS-R.

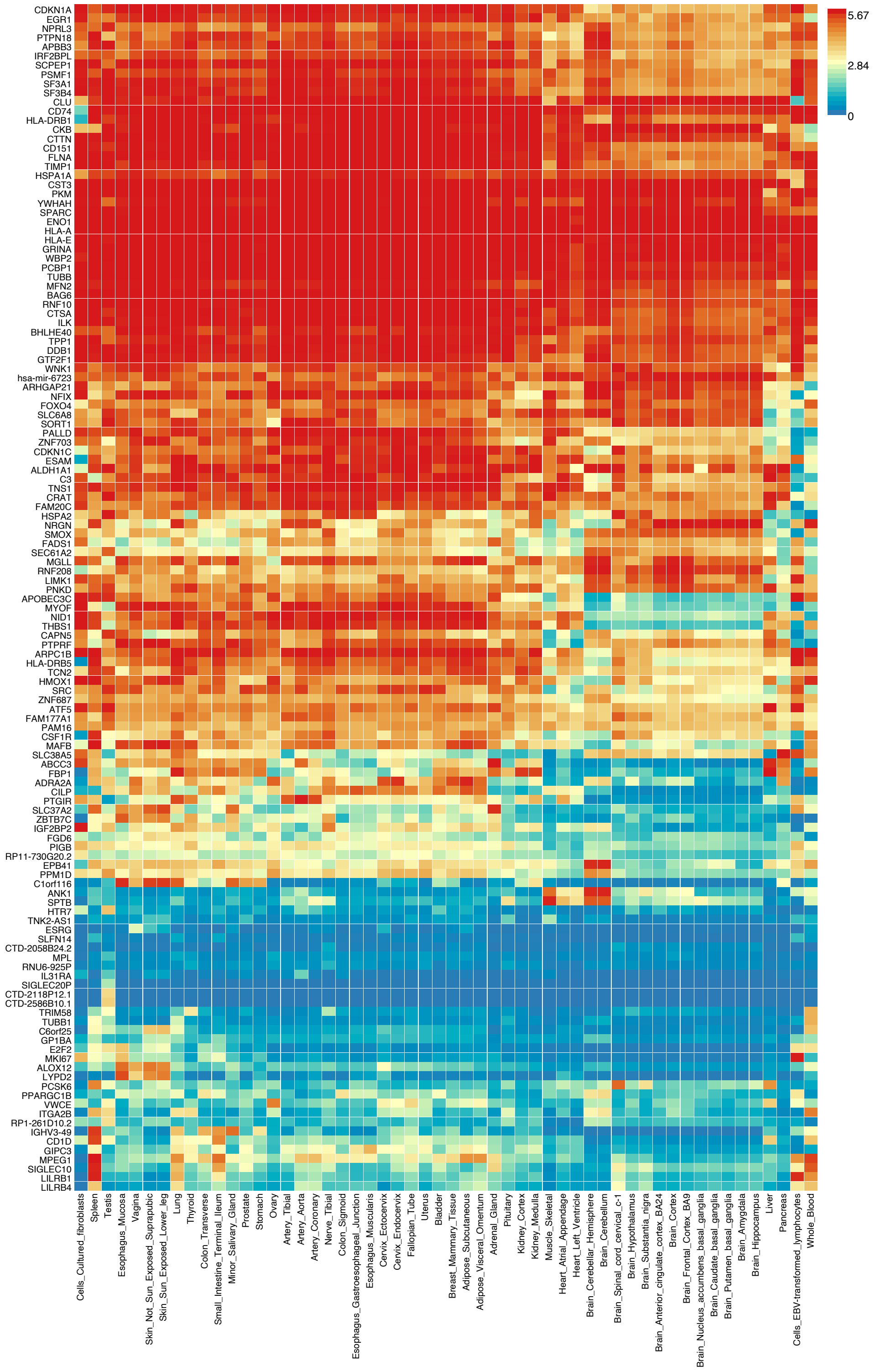


**Supplementary Figure 10**. Genes identified in the longitudinal study (expression vs disease progression). Using tissue expression from GTEx, (n=54, v8) the heatmap illustrating the correlations of expression patterns of 131 recognised genes (of 144) in the longitudinal result (compared to all background). Colour shade indicates average $log_2$ expression level of that gene in corresponding tissue (see legend).

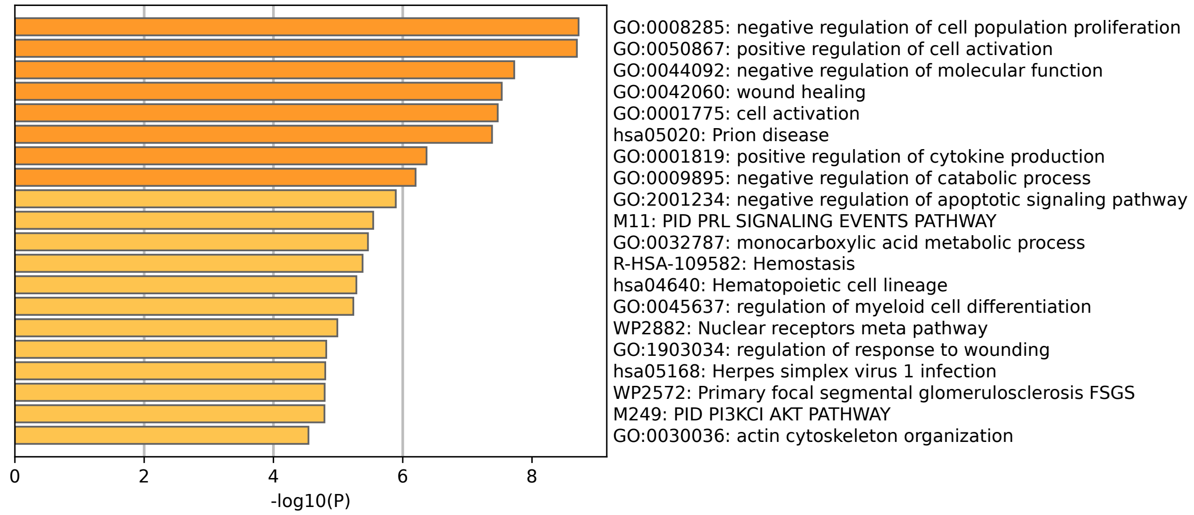


**Supplementary Figure 11**. Result of pathway enrichment analysis on longitudinal result (disease progression vs gene expression) using Metascape. Twenty pathways were enriched.

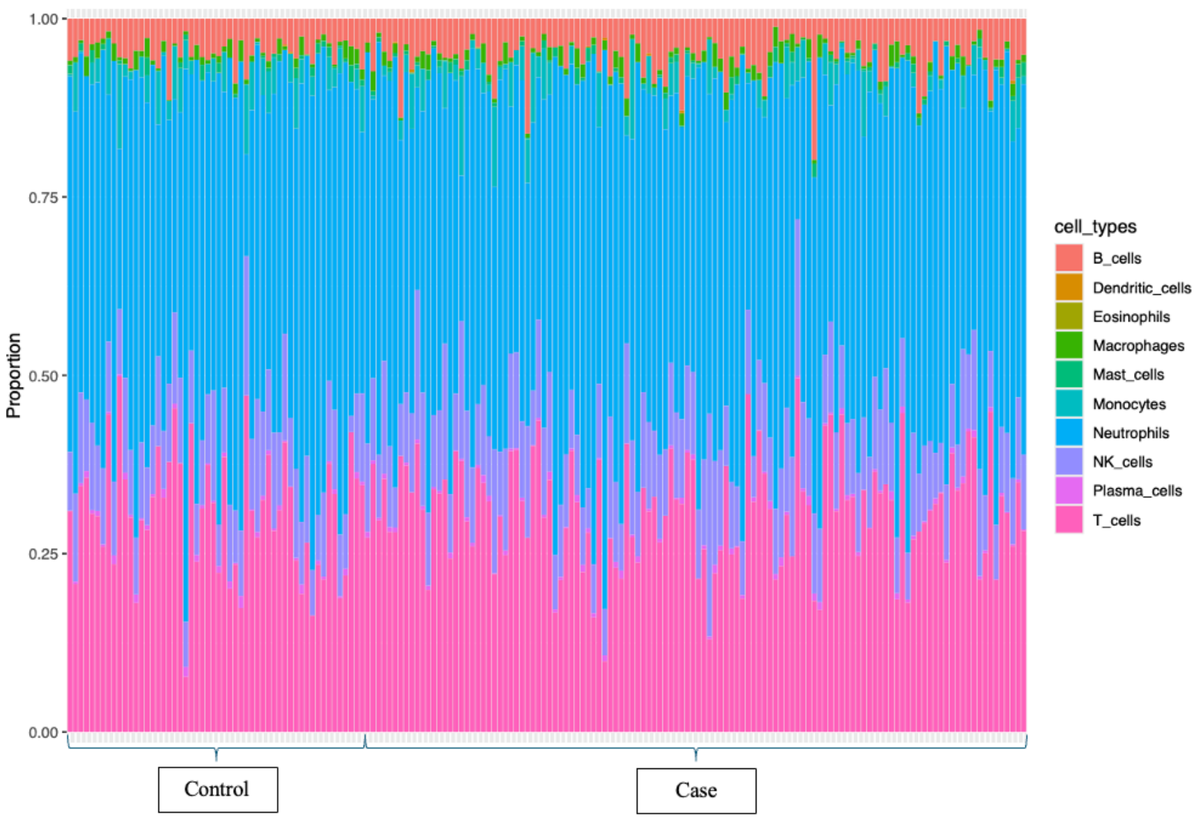


**Supplementary Figure 12.** Estimated cell proportions in the blood samples used for the differentially expressed genes analysis. Each vertical bar represents the cell proportions of a sample after deconvolution. Each colour represents a cell type. Neutrophils accounted for the highest proportion. Eosinophils, plasma cells, macrophages, dendritic cells, and mast cells consisted of only a small proportion.

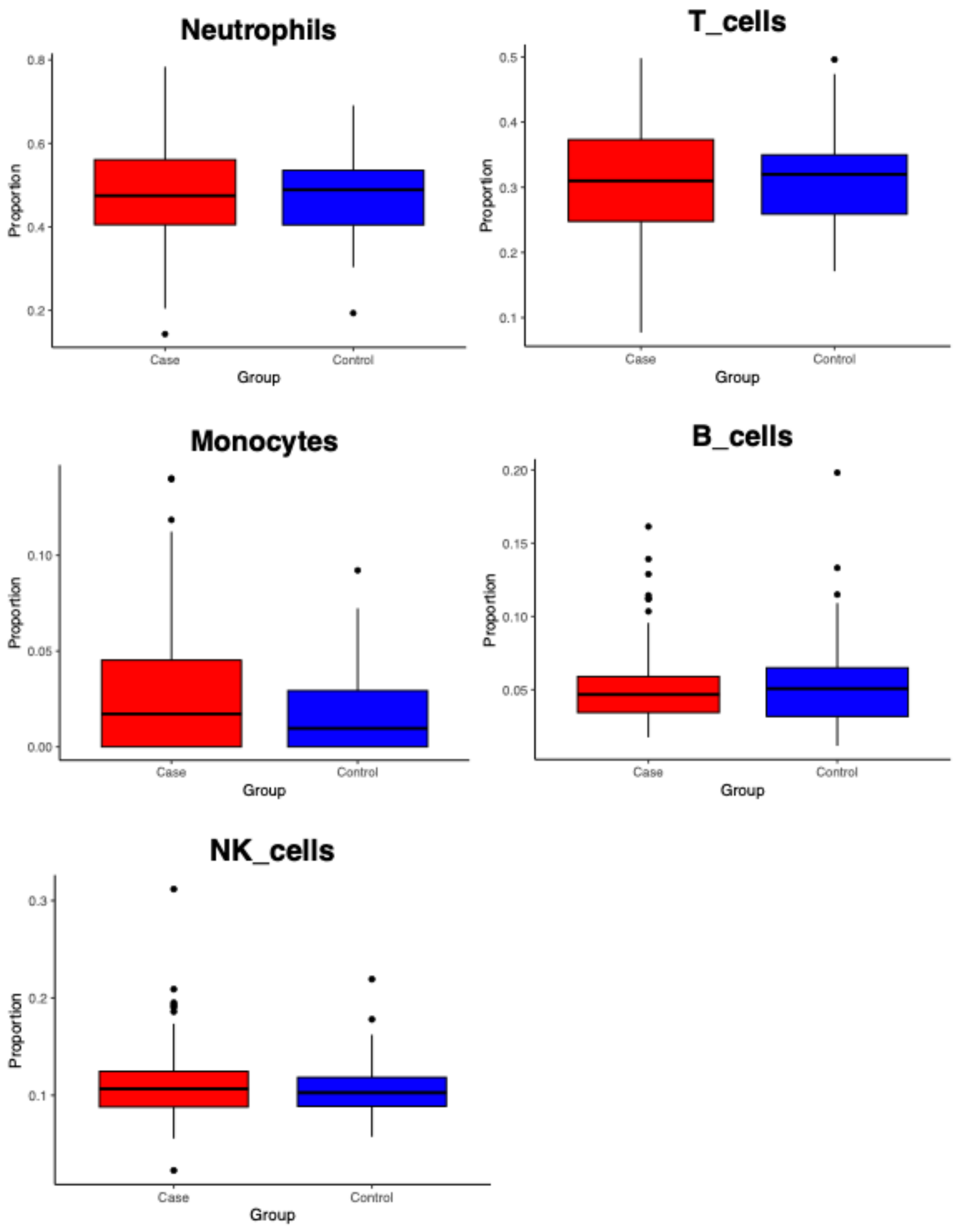


**Supplementary Figure 13.** Proportion of five cell types compared between controls and cases after cell deconvolution using CIBERSORTx. Black dots represent outliers.

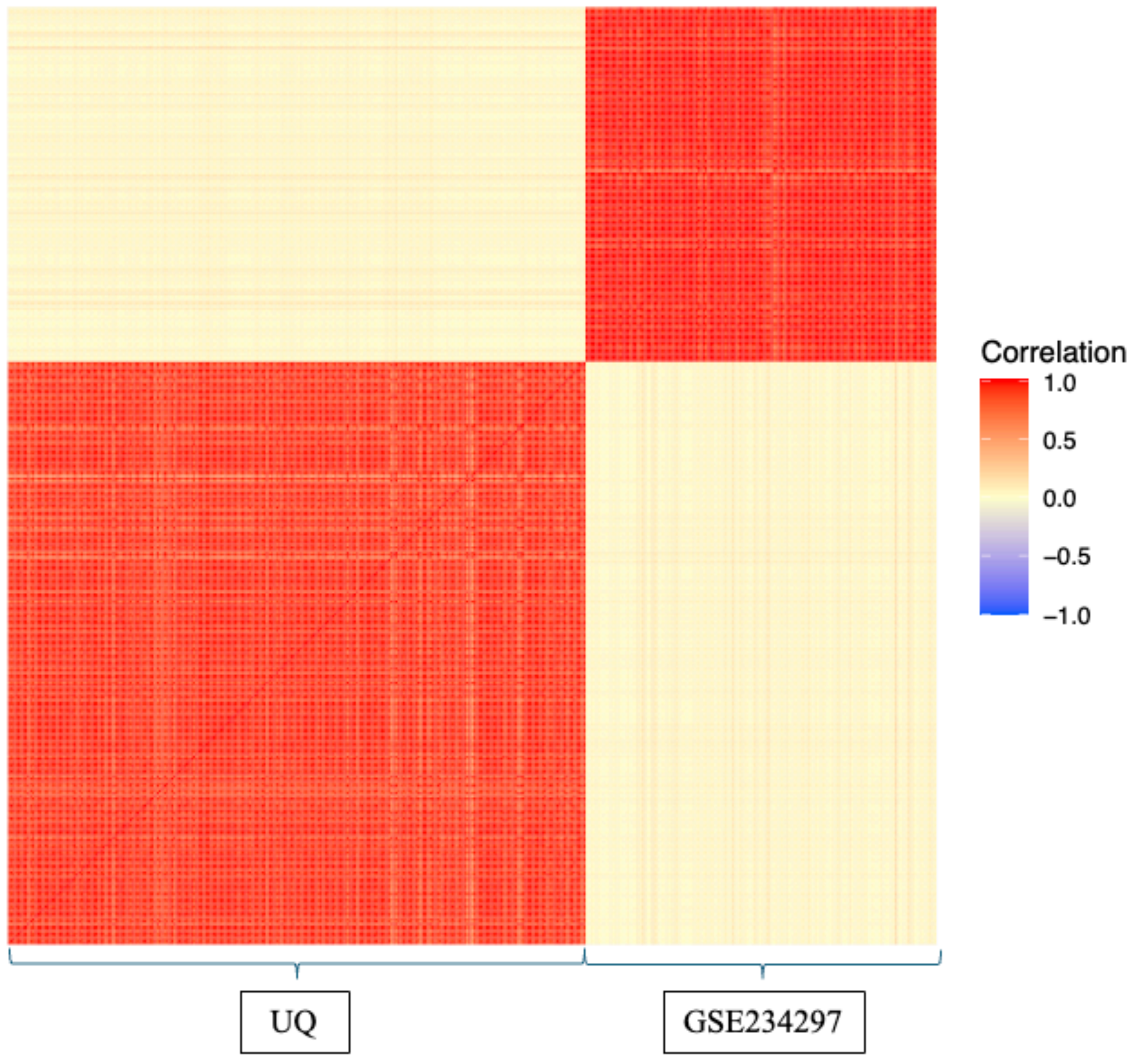


**Supplementary Figure 14**. Heatmap of the correlation matrix between the raw counts of the full dataset in this study and GSE234297. Mean correlation within the pairs of this study was 0.93, GSE234297 = 0.97, UQ vs GSE234297 = 0.52.

**Supplementary Note**

Analysis of blood RNAseq data

The largest publicly available RNA-seq blood dataset in ALS (GSE234974) could not be included in a meta-analysis with our cohort due to differences in processing and lower sequencing coverage. Although both datasets were derived from Australian ALS and control cohorts, using comparable blood collection and extraction protocols (PAXgene tubes with automated extraction), the library preparation methods differed substantially.

The GSE234974 libraries were prepared using the TruSeq Stranded mRNA Sample Prep kit (hereafter TruSeq), while our UQ cohort used the Illumina Stranded Total RNA RiboZero Plus Library Prep kit (hereafter RiboZero). These kits differ in their capture and sequencing profiles. TruSeq enriches for polyadenylated RNA, largely limiting analysis to mature mRNAs, without any Globin/rRNA depletion, while RiboZero depletes ribosomal RNA and captures both polyA and non-polyA transcripts, including non-coding RNAs and pre-mRNAs. Consequently, RiboZero libraries provide greater intron coverage and enhanced detection of alternative splicing events, while TruSeq libraries may exhibit reduced splicing signal due to their 3'-bias and polyA selection.

Additional differences include target sequencing depth (GSE234974: ~50M PE; UQ: ~32M) and quality metrics (e.g., GC bias and lower RIN in GSE234974 vs. higher RIN and no GC bias in UQ data).

To evaluate potential overlap, we re-processed the raw GSE234974 data using our pipeline, omitting the read count filter (≤20M) to retain more samples. This analysis yielded 85 significantly differentially expressed (DE) genes, fewer than the 245 reported in the original study (Grima et al., 2023).

Further comparison revealed limited correlation between the datasets. The Wald statistic values yielded a Pearson correlation of 0.42 ($R^2 = 0.21$), with 67% of all genes falling in the same direction of effect (quadrants I and III).

A correlation matrix heatmap (**Supplementary Figure 14**) showed high within-cohort correlation (UQ: 0.908; GSE234974: 0.975), but very low correlation across cohorts (UQ vs. GSE234974: 0.042), despite all values being positive.
